# A generator-matrix model quantifies the limited contribution of measured biomarkers to human mortality acceleration

**DOI:** 10.64898/2026.07.05.26356402

**Authors:** Masato Tanigawa, Takafumi Iwaki

## Abstract

Aging clocks and biomarkers are increasingly used as if they were the mechanism that drives mortality. But a mechanism, unlike a thermometer, must satisfy three conditions: it must **account** for the mortality signal, be **causal**, and be the **layer that reverses** when aging is reversed. Using public or provider-restricted de-identified data, we test all three. First (accounting), a Markov generator-matrix model with death as an absorbing state, fitted jointly to biomarkers and mortality in NHANES (n=23,512) and replicated in the Health and Retirement Study, assigns most of the Gompertz rise in mortality to a component latent to measured blood biomarkers (92.7%; 89.1% under a mean-field re-specification; 91.5% on replication). Second (causation), a positive-control-calibrated, two-platform cis-pQTL Mendelian-randomization design (UKB-PPP, deCODE) detects known causal proteins (LPA, IL6R) yet finds the measurable inflammatory, renal and growth-signalling markers null. Third (reversibility), at donor level reprogramming reverses a chronological clock (−9.7 and −22.4 yr), whereas a causality-enriched damage clock shows no detectable reversal while somatic identity is retained and moves only as pluripotency is approached. On these data the biomarkers meet none of the three conditions; aging measures appear to track mortality risk rather than its cause, a caution for their use as surrogate end-points.

## Introduction

The last decade produced a proliferation of “aging clocks” and circulating biomarkers of biological age, now entering clinical trials and geroprotector evaluation as putative readouts of the aging process. Their use rests on an implicit premise: that the molecular quantity being measured is, or closely tracks, the *mechanism* that makes us age and die. A thermometer reflects fever but cooling the thermometer does not cure the disease; whether aging biomarkers are mechanism or thermometer is rarely tested directly.

The question is sharpened by three influential and partly competing theories of what drives aging mortality (for a critique of the hallmarks paradigm, see [1]). (A) **Damage accumulation / reliability theory** holds that aging is the macroscopic form of stochastic somatic damage approaching a tolerance threshold; the damage is real and largely irreversible. (B-1) The **hyperfunction** view holds that aging is a quasi-programmed continuation of nutrient/growth (mTOR–insulin/IGF-1) signalling rather than damage [2]. (B-2) The **information theory** of aging holds that aging is reversible loss of epigenetic information, with partial reprogramming restoring youthful states [3,4]. These theories make different predictions about whether the mortality-associated component is measurable, causal, and reversible.

The empirical state of the field leaves the question open. Epigenetic clocks (Horvath [5], Hannum [6], PhenoAge [7], GrimAge [8], DunedinPACE [9]) and proteomic age predictors consistently *predict* mortality and morbidity, and second-generation, mortality-trained clocks do so most strongly; yet prediction is not causation, and the molecular features a clock weights need not lie on the causal path to death. Genetic causal tests of individual aging-associated proteins have been performed piecemeal and give mixed or null results, but rarely within a single framework that calibrates power with positive controls or examines the local genetic architecture by colocalization. In parallel, partial reprogramming reverses epigenetic clocks in vitro and improves some tissues in vivo [4,10,11], fuelling claims that aging is reversible; but whether reprogramming reverses the layer that actually drives mortality, as opposed to a cosmetic chronological tag, has not been separated using clocks that distinguish chronological age from causal damage. What is missing is a test that asks, in one coherent design, three questions at once: how much of mortality acceleration is even captured by accessible measures, whether those measures are causal, and whether they are reversible.

We approach this as a problem in **evaluation science** rather than mechanism discovery: instead of proposing a new aging mechanism, we ask whether the quantities we can already measure are the causal, reversible drivers of mortality, and we adjudicate A / B-1 / B-2 with three orthogonal lines of evidence. First, we quantify how much of mortality acceleration is even *captured* by accessible biomarkers, using a dynamical reliability model (a Markov **generator matrix** with death as an absorbing state) grounded in cross-species reliability theory and the mortality-rate-doubling-time–lifespan scaling recorded across species in AnAge [12,13,14]. Second, we test whether the measurable molecular components are **causal** for human lifespan using cis-pQTL Mendelian randomization plus colocalization on two independent proteomic platforms, anchored by known-causal positive controls so that a null can be bounded against effects of known-causal magnitude. Third, we test **reversibility** by asking whether cellular reprogramming reverses a chronological clock and a causality-enriched damage clock equally.

We find that the three lines converge: the model-inferred mortality-associated component is largely latent to accessible measures, its measurable molecular proxies show no detectable causal effect on lifespan across the inflammatory, renal and growth-signalling axes (extending prior null Mendelian-randomization results for epigenetic clocks [27]), and the causality-enriched damage layer shows no detectable reversal during identity-retaining reprogramming. The popular clocks encode mortality risk, not mechanism.

### Contribution

Individual elements have precedent: phase-type mortality models with biological-aging states [15,16]; piecemeal causal nulls for inflammatory markers [17]; causality-enriched clocks [18]; the broader distinction between the predictive utility of aging clocks and their mechanistic interpretation, recently discussed in perspective [19]; and standardized frameworks for benchmarking the predictive performance of aging biomarkers [20]. The contribution here is not that argument or predictive benchmarking, but its integrated empirical adjudication of cause: a single computational framework that subjects the marker-versus-cause question to three orthogonal, convergent tests: (i) a **generator-matrix reliability model** whose joint biomarker-and-mortality likelihood decomposes mortality acceleration into visible biomarker-driven and latent components; (ii) a **positive-control-calibrated, two-platform cis-pQTL MR layer** (with descriptive colocalization) that tests whether the measurable components are causal; and (iii) a **clock-battery reversibility test** that separates chronological-clock reversal from damage-clock reversal. The specific new causal nulls (renal and growth-signalling axes on parental lifespan), the day-stratified reading of the partial-reprogramming window, and the hierarchical reading of A / B-1 / B-2 are developed in the Discussion.

## Results

The three analyses below are three orthogonal tests of a single thesis: that the molecular quantities we can measure lie largely outside the causal path to mortality. We ask, in turn, *how much* of mortality acceleration is even captured by accessible biomarkers, *whether* the measurable components are causal for lifespan, and *whether* they are reversible. Because each test uses a different methodology, dynamical decomposition, genetic causal inference, and a clock-battery intervention, their convergence is triangulation rather than repetition.

**First condition — accounting: a generator-matrix model shows ∼89–93% of mortality acceleration is latent to the measured blood-biomarker axes**

### How much of mortality is even measurable?

We built a dynamical reliability model that represents the body as accumulating burden across four axes chosen to span the hallmark taxonomy [21] (inflammation, metabolic, organ reserve, haematopoiesis) constructed from the nine PhenoAge blood biomarkers, kept multi-dimensional rather than collapsed to a single index, with death as an absorbing state (Fig. 1). Because the age-at-death distribution of a population constrains the underlying transition dynamics, the model can be fitted jointly to each cohort’s biomarkers and its mortality, and this fit separates the age-related rise in mortality (its Gompertz acceleration) into a *visible* part driven by the measured biomarker axes and a *latent* baseline the measured axes do not capture (full model, equations and priors in Supplementary Methods §1).

**Figure 1.**
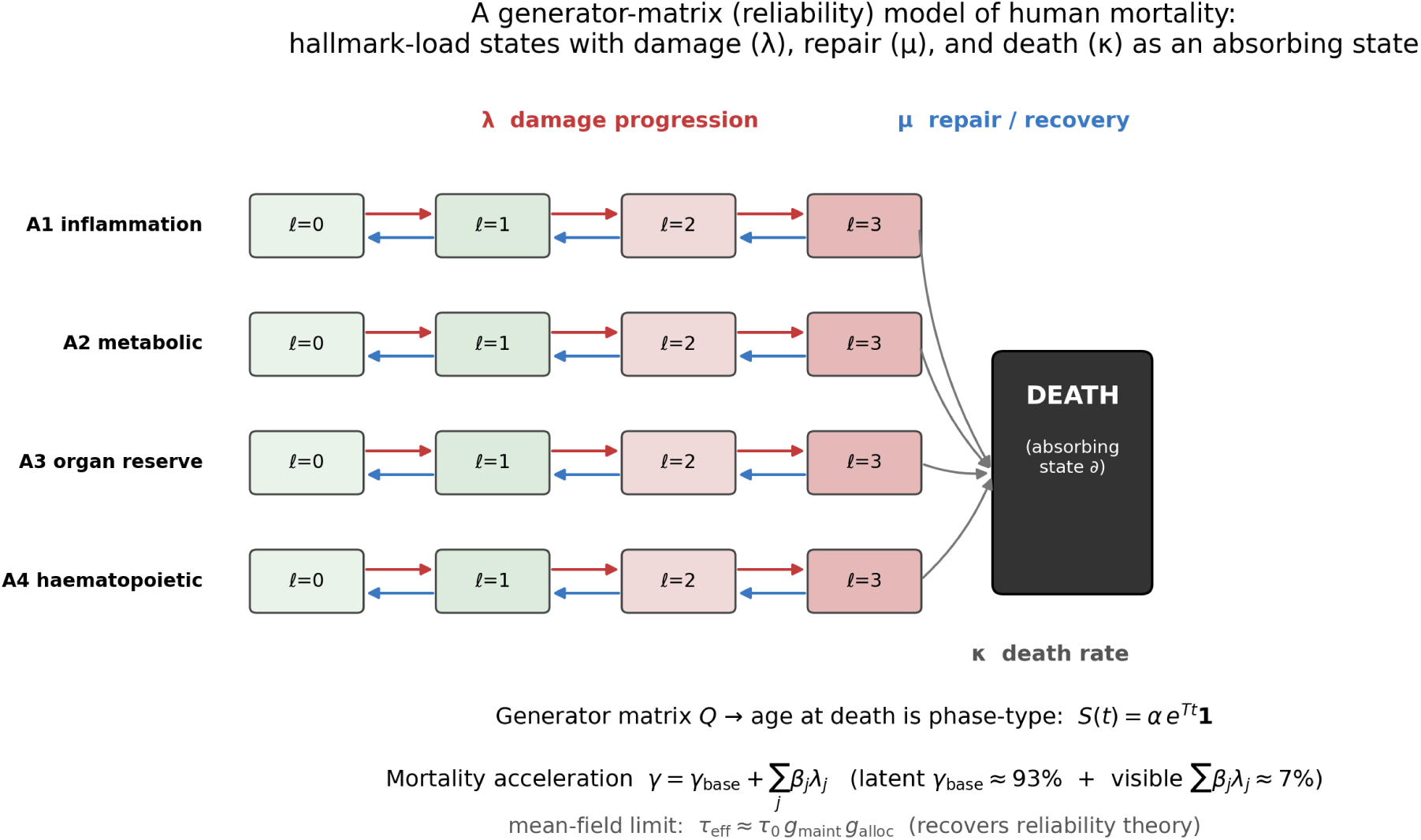
A generator-matrix (reliability) model of human mortality. Hallmark-load states on four axes (A1 inflammation, A2 metabolic, A3 organ reserve, A4 haematopoietic) progress under damage rates and recede under repair rates; death is an absorbing state. The population age-at-death distribution is the resulting phase-type first-passage time, and the Gompertz mortality acceleration decomposes into a latent baseline (∼89–93%) and a visible part driven by the measured axes (∼7–11%); the mean-field limit recovers the reliability decomposition τ_eff ≈ τ0·g_maint·g_alloc (equations in Supplementary Methods §1). (Schematic; source `fig_concept.py`.)

The latent baseline accounts for **∼89–93%** of that rise: **92.7% [92.2–93.2]** in NHANES (1999–2010 pooled, n=23,512, 4,829 deaths) under the primary joint model, **89.1% [88.0–90.0]** under a discretized hallmark-load / mean-field re-specification of the same decomposition, matched to the primary sample and covariates (a 3.6-percentage-point difference; a differently-specified sensitivity analysis rather than an independent first-passage implementation; Supplementary Figure 6, Supplementary Methods §1.3), and **91.5% [89.7–93.3]** on independent replication in a different cohort and period (HRS, prospective mortality) (Table 1; Supplementary Figures 1–2). The measured blood axes thus explain only ∼7–11% of how fast mortality rises with age; among them the haematopoietic and metabolic axes were the largest of a uniformly small set of contributors, an order of magnitude below the latent baseline. The decomposition gave a sharply peaked posterior, was stable to the number of axes and load levels (Supplementary Methods §7), and its structural invariants (that the latent fraction is bounded in [0,1], vanishes when the baseline does, and decreases as the measured axes explain more) were machine-checked in the Lean 4 proof assistant [46] with Mathlib [47] (Supplementary Methods §1.4). A conditional expected-event-count diagnostic, conditioning on each person’s observed follow-up, reproduced the aggregate death count (4,829 observed vs 4,827 expected; with a freely fitted baseline scale this aggregate agreement is close to automatic and is therefore weak evidence of predictive calibration), but revealed residual age-structure misfit: three of six age bands (20–45, 65–75 and 85+) fell outside the 95% replication interval, the largest discrepancy being 65–75 (1,322 observed vs 1,440 expected) (Supplementary Figure 4, Supplementary Table 13). The latent fraction was stable across sex, age group and survey cycle (86.7–95.5%; overall 92.2%; Supplementary Figure 5, Supplementary Table 14), robust to the γ_base prior centre (92.7% across N(0.04–0.12,0.03); Supplementary Table 15), and identified by the joint likelihood in a parameter-recovery test on data simulated from the model itself (unbiased across a 60–95% grid; Supplementary Table 16). The latent is not featureless: added one at a time to the blood axes in HRS, protein-trained DNA-methylation modules for renal (cystatin C), inflammaging and senescence signatures each reduced it, localizing part of the latent to a slowly accumulating, DNAm-integrated systemic load rather than a transcriptional snapshot (Supplementary Results §8.1, Supplementary Figure 3).

**Table 1.**
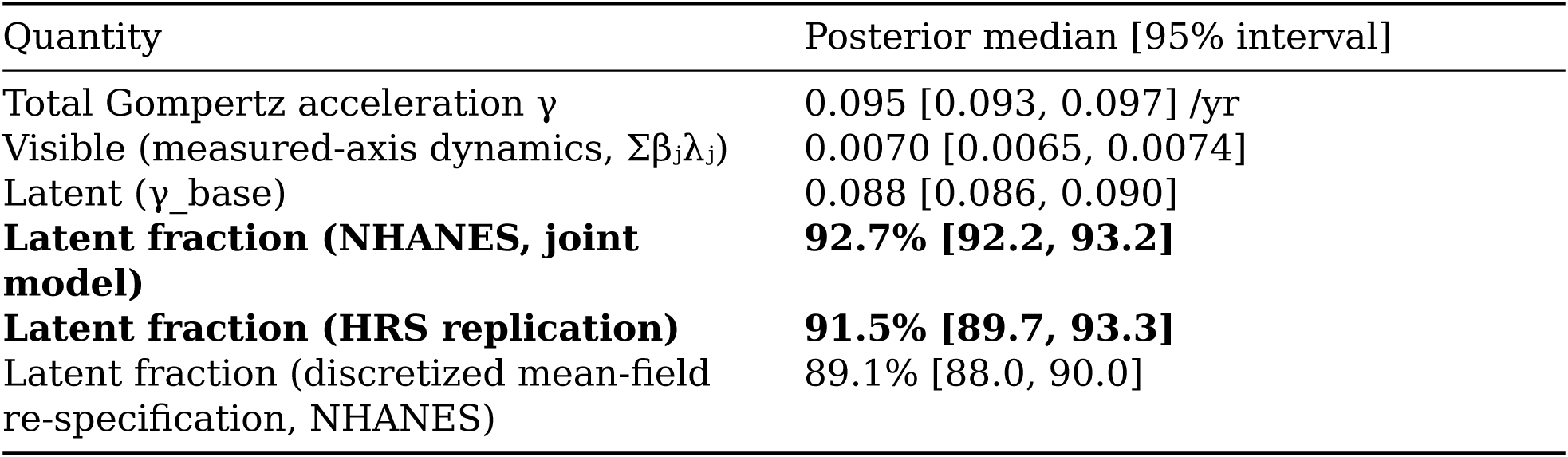
Generator-matrix decomposition of mortality acceleration (NHANES; replicated in HRS). Per-axis contribution (β·λ): A1 inflammation 0.00049; A2 metabolic 0.00202; A3 organ/renal 0.00133; A4 haematopoietic 0.00311. The joint biomarker-mortality model is the primary estimator; a discretized hallmark-load / mean-field re-specification of the same decomposition, matched to the primary complete-case sample and covariates, gives 89.1% (a 3.6-percentage-point difference; a sensitivity analysis, not an independent first-passage implementation). Intervals are 95% highest-density intervals except the HRS replication row, whose posterior draws are summarized as an equal-tailed 95% percentile interval (`hrs_bayes_generator.py`); for these near-symmetric posteriors the two coincide to the precision shown. *Source:* `bayes_unified_summary.csv`, `bayes_unified_axis.csv`, `HRS_bayes_generator.csv`, `phasetype_decomposition_bayes.csv`.

| Quantity | Posterior median [95% interval] |
| --- | --- |
| Total Gompertz acceleration $\gamma$ | 0.095 [0.093, 0.097] /yr |
| Visible (measured-axis dynamics, $\Sigma\beta_j\lambda_j$) | 0.0070 [0.0065, 0.0074] |
| Latent ($\gamma_{\text{base}}$) | 0.088 [0.086, 0.090] |
| **Latent fraction (NHANES, joint model)** | **92.7% [92.2, 93.2]** |
| **Latent fraction (HRS replication)** | **91.5% [89.7, 93.3]** |
| Latent fraction (discretized mean-field re-specification, NHANES) | 89.1% [88.0, 90.0] |

**Second condition — causation: the latent’s measurable components show no detectable causal effect on lifespan (two-platform cis-pQTL MR)**

### Are the measurable markers causal?

If these molecular markers drove mortality, genetically-predicted higher levels should shorten lifespan. We tested this with cis-pQTL Mendelian randomization plus colocalization against parental-lifespan GWAS [22], on **two independent platforms** (UKB-PPP Olink; deCODE SomaScan), with known-causal **positive controls** to calibrate power (methods in Supplementary Methods §2–4). Effects are reported on the outcome’s native scale — SD of the rank-normalized Martingale residual of combined parental attained age per SD higher protein (abbreviated SD/SD); the outcome GWAS is not calibrated in years. Because the UKB-PPP exposure sample (n=54,219) lies within the UK Biobank outcome sample (n=389,166), exposure and outcome overlap; with the strong instruments used here (F=1499–20031) such overlap biases estimates toward the confounded observational association, so a null is the conservative outcome, and the deCODE (Icelandic) exposure arm is free of this overlap (Supplementary Methods §2).

Positive controls behaved as expected and in the biologically correct direction (the parental-lifespan outcome is oriented so that a positive effect lengthens life; Supplementary Methods §2): **LPA** (β=−0.027 SD/SD, p=9×10⁻¹²; harmful) and **IL6R** (β=+0.0086 SD/SD; protective; p=2.8×10⁻⁵ [UKB-PPP], p=3.8×10⁻⁶ [deCODE]) showed clear causal effects, confirming the pipeline detects causation at these instrument strengths and outcome sample sizes. Instruments were strong where proteins could be instrumented (F = 1499–20031); the positive-control effects were precise (LPA β = −0.027 [95% CI −0.035, −0.019] SD/SD; IL6R β = +0.0086 [0.0046, 0.0126]), whereas the latent components’ confidence intervals spanned zero (cystatin C β = −0.0017 [−0.018, 0.014]; GDF15 −0.0033 [−0.019, 0.012]).

For the five primary-platform proteins, results were unchanged across the two MR implementations and the three longevity outcomes tested. By contrast, the latent’s measurable components were **null even with stronger instruments**: GDF15 (F=1924) p=0.68; **cystatin C / CST3** (F=1499) p=0.84, replicated on deCODE (GDF15 p=0.54; CST3 p=0.87). Colocalization is reported descriptively rather than as corroboration, because its sensitivity is not established in this design: neither positive control colocalized (LPA PP.H4 = 0.000 with PP.H3 = 1.000, the expected behaviour where the single-causal-variant assumption fails at an allelically heterogeneous locus; IL6R PP.H4 = 0.345), and over cis-windows of 50–500 kb and p₁₂ priors of 10⁻⁶–10⁻⁴ whether IL6R colocalizes at all is set by the prior (PP.H4 0.047→0.843) while the window is immaterial (≤0.005; Supplementary Table 17). Descriptively, for cystatin C and SERPINE1 the posterior mass sits on exposure-only signal (PP.H1 = 0.785–0.988 and 0.738–0.989 across that grid), that is, the lifespan GWAS shows no detectable signal in these regions; this is a statement about outcome-side power, not about the absence of a causal effect. In particular the nominal SERPINE1 association is not explained by linkage disequilibrium: the distinct-causal-variant posterior never exceeds PP.H3 = 0.082 anywhere in the grid. GDF15 is the one locus at which distinct causal variants are robustly favoured (PP.H3 = 0.589–0.660). B2M and IL6 had no usable cis-pQTL on **either** platform (a biological, not platform-specific, limitation), and TIMP1’s instrument lies on chromosome X, absent from the autosomal lifespan GWAS (Fig. 2; Table 2). The conclusion, that the measurable inflammatory (GDF15) and renal (cystatin C) proxies are consistent with a downstream-marker rather than a causal interpretation, is stable across platform and instrument strength. Formal power analysis makes the nulls quantitative: for the strongly-instrumented markers (GDF15, CST3, IGFBP3 and GHR) the design had ≥80% power to detect an LPA-scale effect (0.027 SD/SD; minimum detectable effect 0.010–0.023 SD/SD), so the nulls among them (GDF15, CST3, IGFBP3) exclude causal effects of that magnitude, while GHR’s nominal association is well powered but does not survive correction; the weakly-instrumented markers (SERPINE1, IGFBP1, IGF1R) are not resolved by this design, and we do not claim to exclude effects as small as IL6R’s (0.009 SD/SD) (Supplementary Table 12; `mr_null_power.py`). The nulls are also robust to the choice of outcome GWAS. Repeating the lead-variant MR against a larger parental-lifespan GWAS (Timmers et al. 2019 [45], GCST009890, ≥500,193 offspring, reported on a log-hazard scale) leaves every non-control marker null and leaves both nominal signals weaker rather than stronger (SERPINE1 p=0.15; GHR p=0.038), although GHR’s direction is the same in both outcome GWAS. This larger GWAS does not, however, add power: the scale-free comparison of outcome-side z-statistics at the same variants gives a geometric mean ratio of 0.91 across the two positive controls, and IL6R is detected less strongly (p=2.8×10⁻⁵ → 0.0055), so Pilling 2017 is retained as the primary outcome and Timmers is reported as a sensitivity analysis only (Supplementary Methods §2; Supplementary Table 18). MR-Steiger directionality supported the protein-to-lifespan direction among the five UKB-PPP instruments tested, each of which explained far more variance in its protein than in lifespan (R²ₑₓₚ/R²ₒᵤₜ = 2.2×10²–2.5×10⁵; all Steiger p < 10⁻²⁷) (`mr_steiger.py`). Across all seven non-control markers tested on the primary platform, the only two nominal associations (SERPINE1, GHR) survive neither Bonferroni correction (α=0.05/7=0.007) nor Benjamini–Hochberg control of the false-discovery rate at 5% (smallest q=0.076), whereas both positive controls survive either; the null is therefore robust to multiple testing under both a conservative and a permissive correction (Supplementary Methods §2; Supplementary Table 18). The growth-signalling/insulin–IGF (hyperfunction, B-1) axis gives the same picture within its limits: the strongly instrumented IGFBP3 is null with ≥80% power against an LPA-scale effect; GHR’s nominal association does not survive correction; IGFBP1 and IGF1R are underpowered; and direct IGF-1 cannot be validly instrumented against this outcome GWAS: its cis lead variant is absent from it, and all three covered genome-wide-significant cis variants are rare (minor-allele frequency 0.37–0.48%), so the estimate they yield is uninformative (β=+0.058 [−0.017, +0.133], p=0.13; minimum detectable effect 0.107 SD/SD, four times the LPA effect; Supplementary Table 18). Prompted by a prior trans-omic Mendelian-randomization study that reported soluble IGF2R causal for parental lifespan, we tested IGF2R on the primary platform as well, outside the pre-specified family: with a very strong instrument (F=2515) it is null (β=+0.0097 [-0.0020,

**Figure 2.**
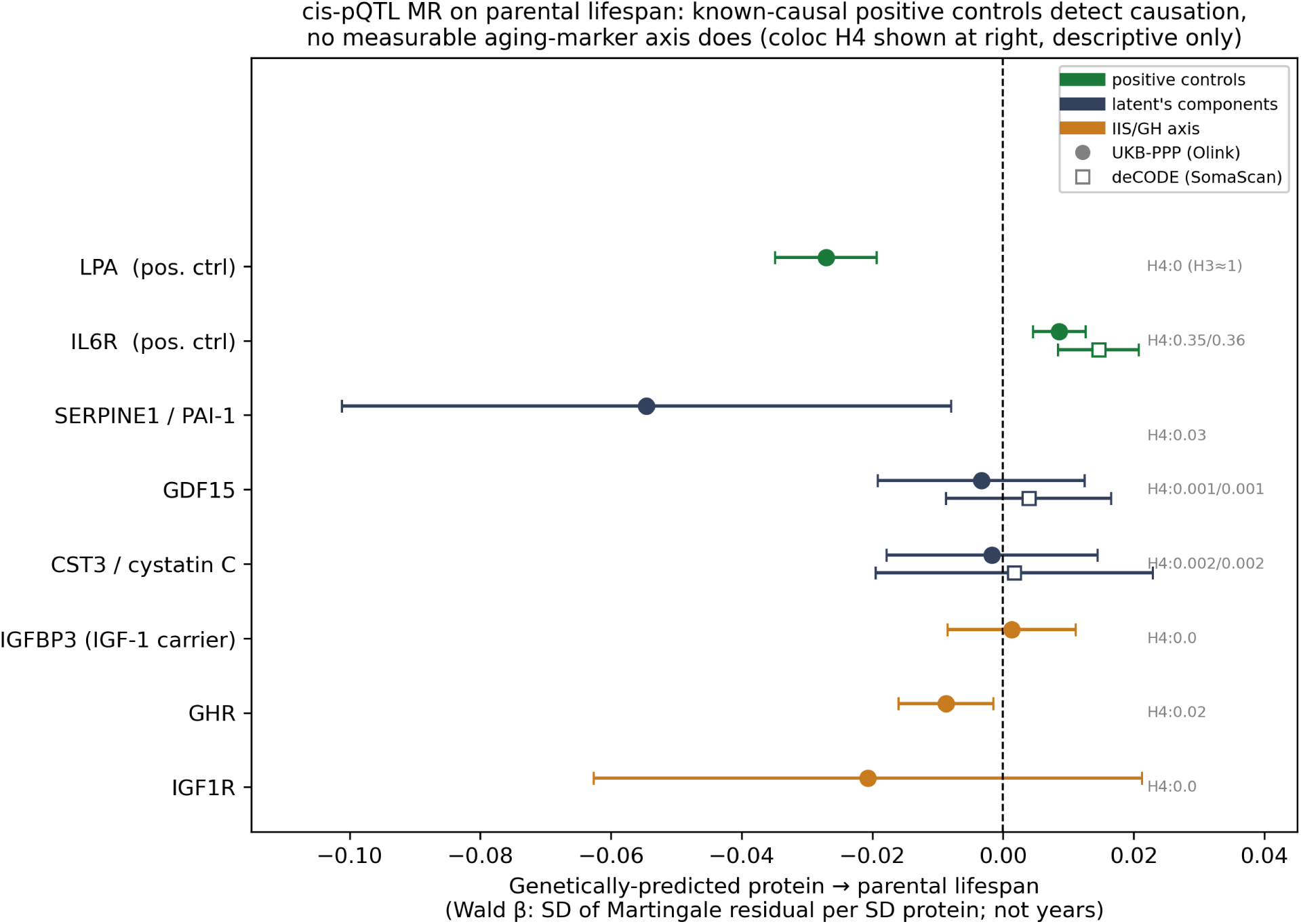
cis-pQTL Mendelian randomization and colocalization of measurable aging-marker axes on human lifespan. Forest plot of genetically-predicted protein effect on parental lifespan (Wald β in SD/SD — SD of the parental-lifespan Martingale residual per SD protein; bars = 95% CI) for known-causal positive controls (green: LPA, IL6R), the latent’s inflammatory/senescence/renal components (dark: SERPINE1, GDF15, CST3/cystatin C), and the growth-signalling/IIS axis (orange: IGFBP3, GHR, IGF1R), on two proteomic platforms (circles UKB-PPP Olink; open squares deCODE SomaScan). Right-hand annotations: colocalization PP.H4 (shared causal variant), shown for description only; because neither positive control colocalized, a low PP.H4 is not interpreted here as evidence against causality (Supplementary Methods §3; Supplementary Table 17). Positive controls (LPA, IL6R) have CIs excluding zero; every measurable molecular component is either null (CI crosses zero) or, where nominally non-zero (SERPINE1, GHR), fails Bonferroni correction (α=0.05/7). (Source: `MR_ukbppp_oriented.csv`, `MR_COLOC_decode_oriented.csv`, `MR_COLOC_iis_oriented.csv`, `COLOC_ukbppp.csv`.)

**Table 2.**
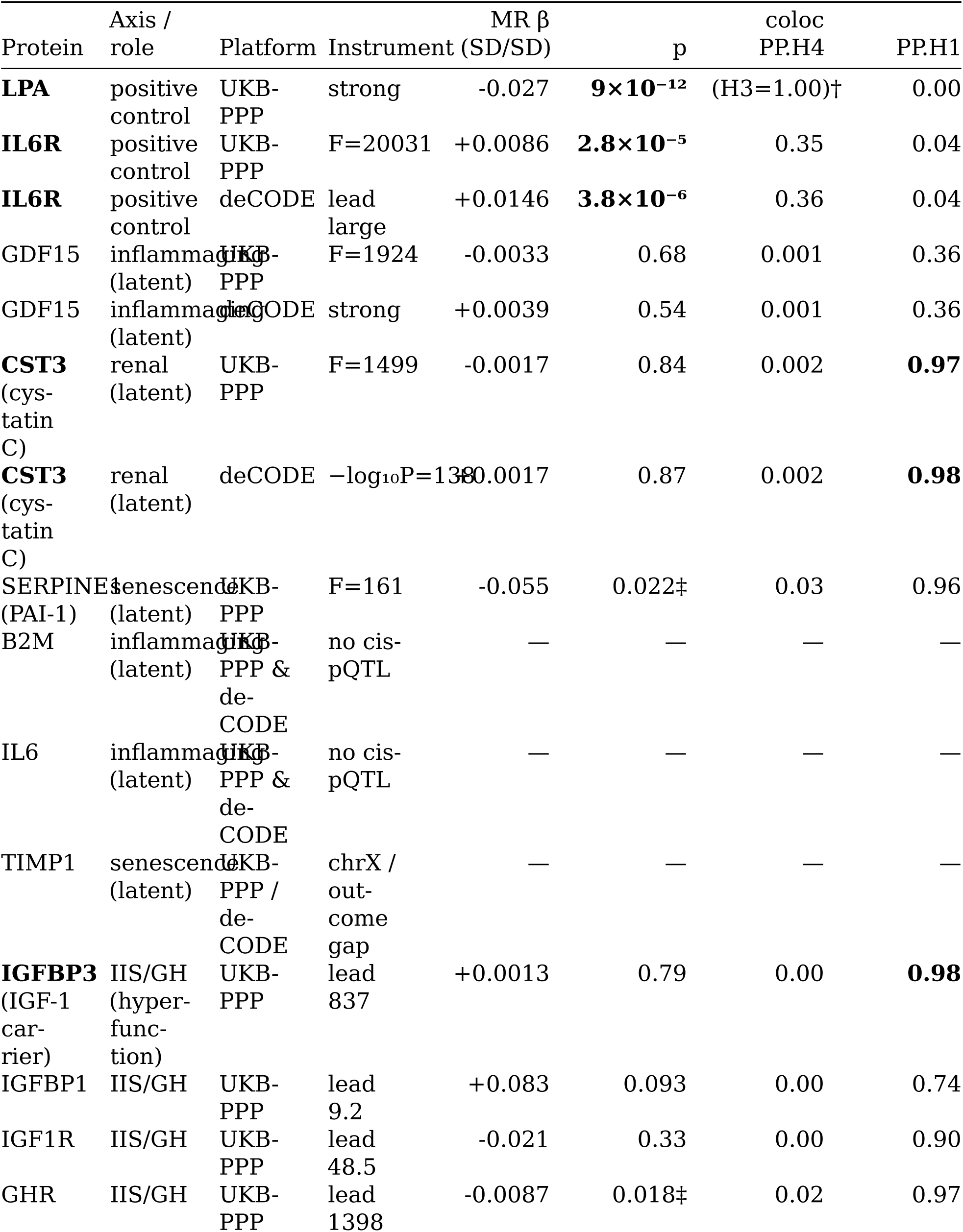
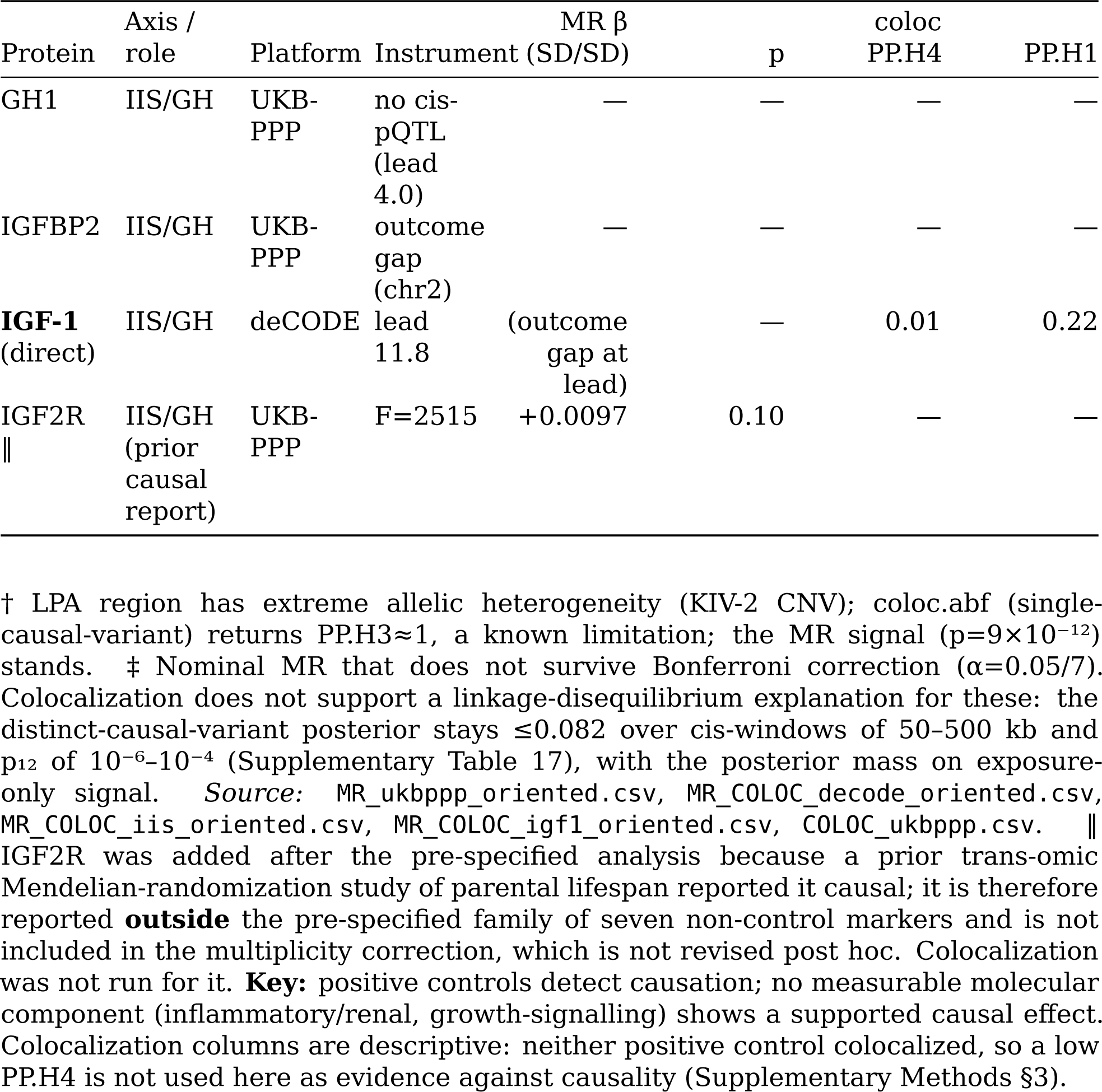
cis-pQTL Mendelian randomization and colocalization of measurable aging-marker axes on human lifespan (Pilling 2017). MR = Wald ratio in SD of the parental-lifespan Martingale residual per SD protein (SD/SD; the outcome is **not** on a years scale); coloc = coloc.abf posterior of a shared causal variant (PP.H4) / exposure-only signal (PP.H1). Platforms: UKB-PPP (Olink), deCODE (SomaScan). Instrument strength = approximate F (UKB-PPP) or cis lead −log₁₀P (deCODE/IIS). “—” = not testable (no usable cis-pQTL, or outcome-GWAS coverage gap).

+0.0213], p=0.10) and the null is informative (minimum detectable effect 0.0167 SD/SD, below the LPA effect), so we do not replicate that report; the two designs differ in proteomic platform (Olink here, SomaScan there, with an exposure GWAS an order of magnitude smaller) and in outcome GWAS. No growth-signalling proxy shows a supported causal effect, so the measurable growth-signalling proxy—like the inflammatory and renal axes—is consistent with a downstream-marker rather than causal role (Supplementary Results §8.2; Fig. 2; Table 2).

**Third condition — reversibility: reprogramming reverses the chronological clock, while the causality-enriched damage clock shows no detectable reversal until somatic identity is lost**

### Is the driver reversible?

Finally we tested reversibility (the core prediction of B-2). To separate a “cosmetic” chronological layer from a mortality-relevant damage layer, we applied a chronological clock (Horvath) and a **causality-enriched, tissue-independent damage clock, DamAge** (derived by Mendelian randomization of CpGs) to cellular reprogramming datasets.

Because the same donors were sampled at several timepoints, all estimates below are **donor-level**: the reprogrammed − control difference is formed within each donor (and day/experiment), averaged over days within donor, and tested as a one-sample t-test on the donor means. In transient reprogramming of human fibroblasts (MPTR [23], 3 donors, 13 matched donor×day×experiment pairs), Horvath reversed (**−9.7**, 95% CI [−18.1, −1.3], p=0.038) while DamAge showed **no detectable reversal** (**+13.1** [−18.0, +44.2], p=0.21; an interval that does not exclude a large reversal), its point estimate sitting in the opposite direction at every timepoint sampled (+13.3, +12.1, +11.5, +17.8 at days 10, 13, 15 and 17; day 15, n=2). In an independent reprogramming method with an internal control (Sendai, SSEA4⁺ reprogramming vs CD13⁺ non-reprogrammed cells; same study; 4 donors, 11 matched donor×day pairs), Horvath again reversed (**−22.4** [−28.0, −16.7], p=0.001) while DamAge showed no detectable reversal (**−2.6** [−7.2, +2.1], p=0.18). Here, however, the behaviour was **day-dependent**: during the partial window neither clock was resolved with four donors (day 9: Horvath −9.8, p=0.16; DamAge +16.6, p=0.19; day 11: −6.2, p=0.45 and +5.8, p=0.54), whereas by day 15, as full iPSC identity is approached, **both** the chronological (−47.2, p=0.001) and the damage clock (−24.4, p=0.030) reversed. The two protocols are therefore consistent with a damage layer that is not detectably moved while somatic identity is retained and moves only as that identity is lost (Fig. 3; Table 3), which places the chronological-versus-damage dissociation of Ying et al. [18] in the partial, identity-retaining window and is compatible with the strong DamAge reversal they reported at the iPSC endpoint. With only three to four donors per protocol these tests have low power: the partial-window contrast is not established at conventional significance, and a non-significant donor-level result must not be read as equivalence. (In a third, independent-group dataset (Sarkar et al. [24], mRNA reprogramming), the reversal was small and normalization-sensitive; our pipeline reproduced the authors’ DNAmAge at r=0.970 per sample, yet the paired reversal differs in magnitude between normalizations (−3.40 yr with the authors’ values, −0.20 yr with ours), so this dataset is an underpowered test of the dissociation; the strongest two-layer evidence rests on the Gill datasets. An in-vivo mouse partial-reprogramming dataset showed only a small, non-significant chronological reversal after age adjustment.)

**Figure 3.**
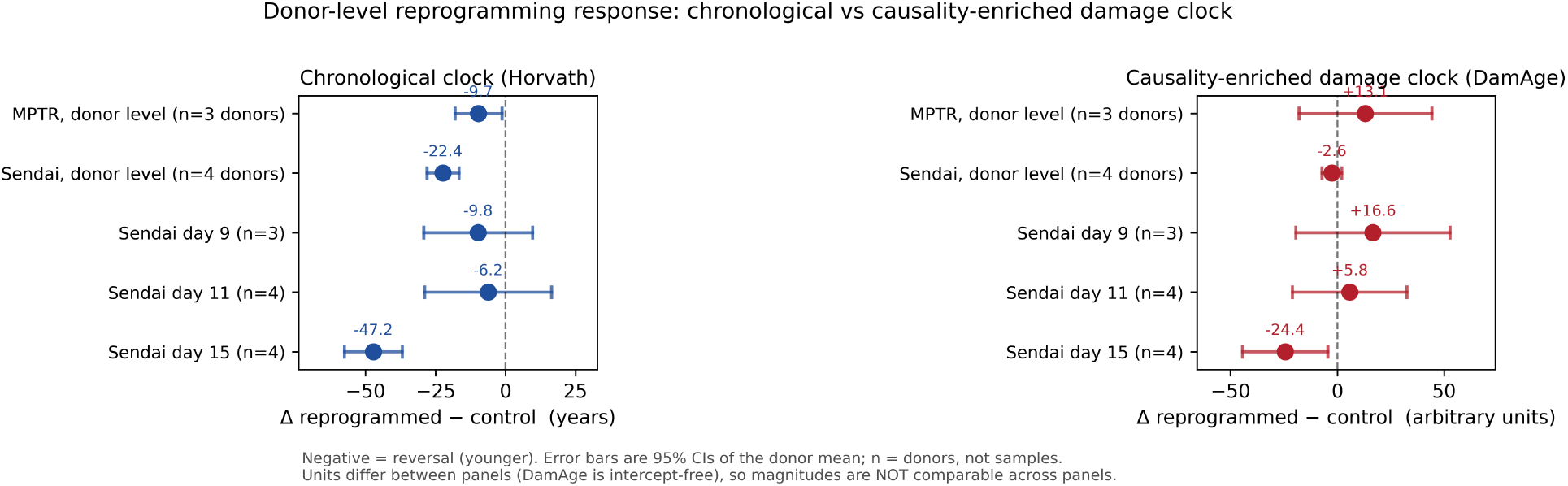
Donor-level reprogramming response of a chronological versus a causality-enriched damage clock. Donor-level Δ (reprogrammed − control; within-donor differences averaged over sampled days/experiments) with 95% confidence intervals, for a chronological clock (Horvath, blue) and a causality-enriched damage clock (DamAge, red), in MPTR (GSE165179; 3 donors, 13 matched pairs) and Sendai (GSE165178; 4 donors, 11 matched pairs). **Units differ between clocks** — Horvath is in years, DamAge is intercept-free and in arbitrary units — so the two are plotted on separate panels and are not directly comparable in magnitude; only the direction and the within-clock comparison across protocols are interpretable. n denotes donors, not samples. GSE142439 is shown for completeness but its public sample identifiers do not resolve donor structure. (Source: `REV_GSE165179_donorlevel.csv`, `REV_GSE165178_donorlevel.csv`, `REV_GSE142439.csv`.)

**Table 3.** Reversibility: donor-level Δ epigenetic age (reprogrammed − control) for a chronological vs a causality-enriched damage clock. Δ is in clock units (years for the age clocks; arbitrary units for DamAge/AdaptAge, which are intercept-free, so only group differences are interpretable); negative = reversal. Each entry is the mean of the per-donor differences (within-donor differences are first averaged over the sampled days/experiments), with a 95% CI and a one-sample t-test on the donor means. **Day stratification.** MPTR is day-stable (DamAge +13.3 / +12.1 / +11.5 / +17.8 at days 10/13/15/17). Sendai is day-dependent: Horvath −9.8 (d9, p=0.16), −6.2 (d11, p=0.45), −47.2 (d15, p=0.001); DamAge +16.6 (d9, p=0.19), +5.8 (d11, p=0.54), −24.4 (d15, p=0.030) — the damage clock moves only as full iPSC identity is approached. These donor-level estimates supersede an earlier pooled comparison that treated repeated measurements of the same donors as independent. With 3–4 donors per protocol the tests have low power; a non-significant result is not evidence of equivalence. § GSE142439 reversal is small and normalization-sensitive: over all eight donor pairs the authors’ own DNAmAge gives −3.40 yr (p=0.023) while our noob processing gives −0.20 yr (p=0.78), the two agreeing closely per sample (r=0.970) but not in the within-pair difference (`rev_gse142439_author_check.py`) and its public sample identifiers do not resolve donor structure, so it is reported as an underpowered, non-donor-level test of the dissociation. In-vivo mouse partial reprogramming (Browder, GSE190665): chronological mammalian clock −0.09 yr (age-adjusted, NS). *Source:* `REV_GSE165179_donorlevel.csv`, `REV_GSE165178_donorlevel.csv`, `REV_GSE142439.csv`, `TRACK2_mouse_clock_ageadj.csv`. **Key:** at donor level the chronological clock reverses in both Gill protocols, while the causality-enriched damage clock shows no detectable reversal while somatic identity is retained, moving only as iPSC identity is approached.

| Clock | Type | MPTR<br>(GSE165179), 3<br>donors | Sendai<br>(GSE165178), 4<br>donors | mRNA<br>(GSE142439) § |
| --- | --- | --- | --- | --- |
| **Horvath2013** | chronological | **-9.7 [-18.1,**<br>**-1.3], p=0.038** | **-22.4 [-28.0,**<br>**-16.7],**<br>**p=0.001** | -0.20 |
| PhenoAge | phenotype/mortality | -7.0 [-50.3,<br>+36.3], p=0.56 | -13.8 [-17.1,<br>-10.6], p=0.001 | +3.5 |
| **DamAge** | causality-enriched<br>damage<br>(MR-derived) | **+13.1 [-18.0,**<br>**+44.2], p=0.21** | **-2.6 [-7.2,**<br>**+2.1], p=0.18** | +1.7 |
| AdaptAge | causal-<br>adaptive | -12.7 [-31.0,<br>+5.5], p=0.095 | -17.0 [-28.4,<br>-5.6], p=0.018 | -3.5 |

## Discussion

The three analyses are not independent checks but one escalating interrogation of a single question — is the measurable the mechanism? A driver of mortality must satisfy three necessary conditions: it must **account** for the mortality signal, be **causal**, and be the **layer that reverses** when aging is reversed. On these data the measurable biomarkers meet none. The model-inferred mortality-associated component of human aging (i) is **largely latent** to accessible biomarkers, so measured markers do not account for it (within the generator-matrix decomposition, ∼89–93% across specifications); (ii) is **not** detectably the measurable molecular markers (across the inflammatory/renal and growth-signalling axes no measurable proxy shows a supported causal effect while known-causal positive controls are detected, extending prior null Mendelian-randomization results for epigenetic clocks [27]); and (iii) in its causality-enriched damage-clock proxy shows **no detectable reversal during identity-retaining reprogramming**, unlike the reversible chronological tag. The convergence across three increasingly stringent conditions, not any single test, carries the conclusion.

### A two-layer model

Reprogramming, and by extension the popular discourse of “rejuvenation,” moves a **reversible chronological tag** (Horvath; not mortality-associated in HRS, HR 1.03, NS) but leaves a **causality-enriched damage layer** (DamAge; which we propose corresponds to the latent) without detectable movement during the partial reprogramming compatible with retained cell identity. This dissociation reframes what aging clocks measure: the popular clocks are best read as **encoders of mortality risk** (or chronological tags) rather than as measures of the causal mechanism, in agreement with the observation that mortality-trained composites capture the latent (circularly) while generic chronological clocks do not.

### Adjudicating A / B-1 / B-2

The reversible information layer (B-2) demonstrably exists, but it is the *non-lethal* chronological layer; the causality-enriched damage layer is, on these data, not detectably reversed, favouring the damage/reliability base (A). Furthermore, the hyperfunction axis (B-1) shows, in our hands, **no supported causal effect in either of its two strongly instrumented circulating proxies**: IGFBP3 is null with ≥80% power against an LPA-scale effect, and GHR’s nominal association survives neither correction. GHR deserves naming rather than aggregating, because it is the one marker in this study with a sub-threshold signal and its direction is the one the growth-hormone literature predicts: plasma GHR is the shed soluble ectodomain, which is low in growth-hormone insensitivity, so higher levels plausibly index greater receptor abundance and hence more signalling, and we estimate higher levels to shorten life. Three things keep this from being a finding. It survives neither Bonferroni nor false-discovery-rate control; it is unreplicated, in that the GHR variant reported for human longevity (rs4130113) lies 111 kb from our instrument and shows no parental-lifespan association in this outcome GWAS (p=0.73); and the premise that soluble GHR tracks tissue receptor abundance is itself postulated rather than established. We therefore report it as the axis’s open question rather than as support for or against B-1. The remaining proxies are unresolved or untestable; like A’s damage and B-2’s reversible tag, its mechanism (if any) is not visible in accessible measures. The unifying theme is therefore not a contest between molecular theories but a **hierarchy**: a damage/reliability base that carries the mortality acceleration but is latent to current measures, beneath a reversible, non-lethal molecular surface (chronological tags, circulating growth-signalling proxies) that is what assays and reprogramming actually move.

### Comparison with prior evidence

The individual nulls are each in line with scattered prior reports, which our framework unifies. The cystatin C null accords with its status as a renal-filtration marker rather than a causal longevity factor; circulating CRP has repeatedly failed Mendelian-randomization tests for the outcomes it predicts, and a recent Mendelian-randomization study independently found CRP and GDF15 non-causal for all-cause mortality while IL6 and IL6R were causal [17]; and human IGF-1/insulin–IGF-signalling MR has been inconsistent despite the strong model-organism phenotypes, which act on intracellular signalling that secreted-protein instruments do not capture. Rather than treating these as isolated negative findings, the positive-control-calibrated, two-platform design shows them to be a single regularity: measurable molecular age markers, across inflammatory, renal and growth-signalling axes, show no detectable causal effect on lifespan, consistent with lying off the causal path to death. That the same class of methods (proteome-wide cis-pQTL MR with colocalization) recovers causal protein effects on human longevity-related outcomes for other proteins [25], and that our positive-control-calibrated power analysis shows the nulls for the strongly instrumented markers are not attributable to low power against effects of LPA magnitude, together indicate that the consistent null across these aging-marker axes is not a detection failure; our design extends the recent all-cause-mortality nulls to parental lifespan and to the renal (cystatin C) and growth-signalling axes. The reversibility result builds on Ying and colleagues’ causality-enriched clocks [18], which uncoupled damage from adaptation and showed DamAge reversing at full iPSC reprogramming: in two independent reprogramming methods we find that within the partial, identity-retaining window the causality-enriched damage layer shows no detectable reversal, moving only as full iPSC identity loss is approached, and we propose that this layer corresponds to the within-human latent, a correspondence argued from their shared properties rather than measured directly. Finally, the picture is compatible with the view that aging clocks track accumulating stochastic variation read out as a clock [26]: a quantity that predicts mortality, can be partially reset, and yet is not the mechanistic driver. Consistent with this, recent Mendelian-randomization work found no evidence that commonly used epigenetic clocks or telomere length causally increased lifespan [27]; the present analysis extends that conclusion from composite clocks to specific protein axes with two-platform replication and positive-control calibration, and integrates it with the latent-fraction and reversibility tests. What the latent damage is (accumulated tissue injury, genomic instability, or a senescent-cell burden poorly represented in circulating blood) remains the central open question.

### Implications

(1) For biomarker-guided geroscience: a measured aging clock or protein shifting under an intervention does not establish that the intervention touched the mortality-associated component; causal (Mendelian-randomization) and reversibility tests of the kind used here should gate such claims. (2) For reprogramming: the chronological reversal that headlines “rejuvenation” is, on these data, the non-lethal layer; demonstrating reversal of the causality-enriched damage layer in vivo (with retained somatic identity) remains the key unmet test. (3) For mechanism discovery: the next task is to **co-develop measures that capture the damage substrate itself**, beginning from the DNAm inflammaging/senescence/renal signatures that partially index it, at tissue and single-cell resolution. More broadly, the framework is reusable: any candidate aging biomarker or geroprotector readout can be passed through the same three gates (latent-fraction accounting, positive-control-calibrated causal MR, with colocalization only where its sensitivity is demonstrated, and chronological-versus-damage reversibility) before it is treated as mechanism. Applying such gates would temper over-interpretation of clock movement in intervention trials and would redirect target discovery from the abundant, measurable surface that shows no detectable causal effect toward the latent damage substrate itself.

### Limitations

The visible/latent decomposition is model-dependent (four linear axes + proportional Gompertz; replicated across two cohorts but not assumption-free), and the identity of the latent (which unmeasured damage) remains unknown. Two caveats bound how the latent fraction should be read. First, the per-axis drift is estimated cross-sectionally; because selective survival tends to flatten cross-sectional age gradients relative to within-individual change, the visible component is plausibly a lower bound and the latent fraction (89–93%) is best interpreted as an upper bound, to be tightened by longitudinal biomarker trajectories. Measurement error in the biomarkers pushes in the same direction and is not modelled here: single-occasion assays of quantities with substantial within-person variability (C-reactive protein above all) attenuate the estimated per-axis coefficients, shrinking the visible part and inflating γ_base, so this is a second and independent reason to read the latent fraction as an upper bound. More generally γ_base is a residual: it is whatever acceleration the measured axes do not account for, and its magnitude could in principle reflect unobserved heterogeneity in individual frailty [34] or misspecification of the proportional-Gompertz form as well as unmeasured damage. Our reading of it as damage-like rests on its behaviour (it is partially indexed by DNAm damage signatures and it resists reprogramming), not on the decomposition alone. Second, the fraction is defined relative to the accessible blood-biomarker panel, quantifying what these particular measured axes fail to capture rather than an absolute unmeasurability: a mortality-trained composite (GrimAge) captured roughly two-thirds of the latent (see the DNA-methylation localization result; Supplementary Results §8.1), so richer or mortality-optimized panels would shrink it. Third, the model decomposes the *rate* structure of the mortality acceleration rather than serving as a calibrated absolute-risk predictor: it reproduces the aggregate and mid-life death counts closely but, as a parsimonious linear-in-age Gompertz form, over-smooths the age-specific death curve: three of six age bands (20–45, 65–75, 85+) lie outside the 95% replication interval, so the misfit is not confined to the extremes. NHANES age top-coding also differs between cycles — 85 in the 1999–2006 cycles but 80 in the 2007–2010 cycles — so the oldest band mixes differently-censored ages (Supplementary Figure 4). The subgroup analysis nonetheless shows the latent fraction is not an artefact of any one sex, age band or survey wave. MR is constrained by instrument availability: B2M and IL6 lack usable cis-pQTL on both platforms, TIMP1 and IGFBP2 fall in outcome-GWAS gaps (chromosome X; a chr2 coverage gap), and coloc.abf assumes a single causal variant per region (limiting interpretation at allelically heterogeneous loci such as LPA). More fundamentally, colocalization has no demonstrated sensitivity in this design (neither positive control colocalized, and the IL6R verdict is set by the p₁₂ prior), so it is reported descriptively and no conclusion rests on it. Circulating pQTL-MR indexes secreted proteins, not intracellular signalling (mTOR), so B-1 is engaged at its measurable proxy, not its mechanism. Aptamer- and antibody-based proteomic platforms can disagree for the same protein, so our failure to replicate the prior SomaScan-based IGF2R result bounds that result on the Olink platform rather than refuting it outright. Reversibility evidence is predominantly in vitro; the independent-group replication was small and normalization-sensitive, and no mouse causality-enriched damage clock exists to test the damage layer in vivo. With three to four donors per protocol, the absence of a detectable DamAge reversal is not a demonstration of resistance, and the correspondence we draw between the damage clock and the within-human latent is inferential (shared properties), not measured. The causal null for the epigenetic axis is drawn from prior Mendelian-randomization work [27], not tested here. DamAge is derived from blood EWAS (pan-tissue but not strictly tissue-independent). A further caveat is that, because DamAge weights CpGs selected by Mendelian randomization, its resistance during partial reprogramming could in part reflect the clock’s construction (if the causality-enriched CpGs happen not to be reprogramming targets) rather than biological irreversibility alone; disentangling these would require testing the overlap between the DamAge CpG set and the reprogramming-responsive CpGs. Because Timmers 2019 contains the UK Biobank sample analysed by Pilling 2017, the outcome-GWAS sensitivity analysis is a check on robustness, not an independent replication, and its weaker detection of the IL6R positive control is itself a reason not to adopt it as the primary outcome. HRS follow-up is short; mortality-trained clocks are intrinsically circular. All conclusions concern public de-identified data and are framed in the project’s ethics stance (social-determinant-first interpretation; no genetic determinism).

## Materials and methods

### Unified inference framework

All three analyses share a single Bayesian causal/graphical-inference framework: the generator-matrix model is Bayesian inference over a continuous-time probabilistic graphical (Markov) model with an absorbing state; Mendelian randomization is causal-directed-acyclic-graph instrument design (variant → exposure → outcome); and colocalization (coloc.abf) is Bayesian model comparison over causal configurations (H0– H4). The marker-versus-cause question is thereby treated as one of Bayesian causal inference.

### Data and cohorts

#### NHANES + Linked Mortality

We pooled the U.S. National Health and Nutrition Examination Survey continuous cycles 1999–2010 [28] (n = 23,844 adults aged ≥ 20 with the nine PhenoAge clinical biomarkers and Public-Use Linked Mortality Files follow-up to 2019, deaths = 5,005; the covariate-adjusted primary model uses the 23,512 complete cases with sex, BMI and smoking, deaths = 4,829). Each biomarker was z-standardized to the pooled 1999–2010 mean and SD (protective markers sign-flipped) and averaged into its axis; robustness to inter-cycle assay differences is shown by the stability of the latent fraction across survey cycles (Supplementary Table 14). **HRS.** Replication used the Health and Retirement Study [29] (RAND HRS Longitudinal mortality × Venous Blood Study biomarkers × HRS DNA-methylation clocks), linked by respondent identifier and analysed locally in aggregate (cell sizes ≥ 5) under the HRS data-use terms; analytic n = 3,985, deaths = 725 over a mean 5.7-year follow-up. **GEO** reprogramming methylation: GSE165179 (MPTR), GSE165178 (Sendai), GSE142439 (mRNA/ERA), GSE247179 (chemical), GSE190665 (in-vivo mouse 4F [10]). **pQTL:** UKB-PPP Olink (Synapse syn51365303; [30]) and deCODE SomaScan [31]. **Outcome GWAS:** parental lifespan (GWAS Catalog [32]

GCST006697, harmonised GRCh38; Pilling et al. 2017 [22]) — combined parental attained age as rank-normalized Martingale residuals in n = 389,166 British-ancestry UK Biobank participants, i.e. **not** a years scale; parental top-decile longevity (GCST006698; Pilling et al. 2017) and parental extreme longevity (GCST003395; Pilling et al. 2016 [33]) were used as secondary outcomes where available. A larger parental-lifespan GWAS (Timmers et al. 2019 [45], GCST009890, ≥500,193 offspring, UK Biobank combined with the LifeGen consortium, effects reported as log hazard protection ratios) was used only for the outcome-GWAS sensitivity analysis; it is released on GRCh37 with no harmonised build, and UKB-PPP carries no rsIDs, so variants were bridged GRCh38→rsID→GRCh37 through the harmonised GCST006697 file rather than lifted over (`outcome_gwas_comparison.py`). **Cross-species:** AnAge [13].

### Generator-matrix model of mortality acceleration

We represented the organism as occupying discrete hallmark-load states on four biomarker axes (built from the nine PhenoAge NHANES biomarkers, kept multi-dimensional rather than collapsed to one index; the mortality model is adjusted for sex, BMI and smoking status (ever/never), never race or nationality), with death as an absorbing state, so that the population age-at-death distribution is a phase-type first-passage time. A joint Bayesian likelihood ties the state dynamics (per-axis drift) to a proportional-hazard Gompertz mortality; this breaks the identifiability of mortality-only phase-type fits [15,16] and decomposes the Gompertz acceleration into a *visible* biomarker-driven part and a *latent* baseline, the latent fraction being the baseline’s share (`bayes_unified_generator.py`). The primary estimate is this joint proportional-hazard model; a discretized hallmark-load / mean-field re-specification of the same decomposition (matched sample and covariates) provides a sensitivity cross-check rather than an independent first-passage implementation, and the decomposition’s structural invariants (bounds, sign, limits) were machine-checked in Lean 4 [46] with Mathlib [47]. The baseline prior is anchored by the cross-species mortality-rate-doubling-time–lifespan relation, connecting the model to reliability theory. Full state space and generator, equations, priors, sampling, both estimators, and the formal verification are given in Supplementary Methods §1, with prior- and discretization-sensitivity in §7.

### DNAm clock battery and latent localization

Epigenetic clocks (Horvath, Hannum, PhenoAge, and the Ying et al. [18] Dam-Age/AdaptAge/CausAge causal clocks; the universal mammalian clock [40] for mouse) were scored with a streaming pure-Python engine (`clocks.py`). To localize the latent, protein-trained DNAm surrogate modules (inflammaging, senescence, metabolic, renal) were added one at a time to the four blood axes in HRS and the reduction in latent fraction recorded; GrimAge, DunedinPACE and whole-blood RNA-seq inflammaging/senescence gene modules were Cox-modelled for mortality in parallel. Full module definitions, imputation and coverage handling are in Supplementary Methods §5.

### Mendelian randomization

Two-sample cis-pQTL Mendelian randomization [41] used genome-wide-significant cis variants (±1 Mb) from UKB-PPP (Olink) and deCODE (SomaScan) as instruments and the parental-lifespan GWAS [22] as outcome (via the MRC IEU OpenGWAS API [42] or the harmonised GWAS-Catalog file). Effects were harmonized to the protein-increasing allele and the Wald ratio (SD of the outcome per SD protein; SD/SD) taken as the causal estimate (inverse-variance-weighted with MR-Egger/weighted-median sensitivity where multiply instrumented). Exposures were the latent’s measurable components and the growth-signalling/IIS axis; **known-causal positive controls (LPA, IL6R)** were analysed identically to calibrate power, and instrument strength summarized by the F-statistic. Full instrument construction and estimators are in Supplementary Methods §2, and the minimum-detectable-effect power analysis in Supplementary Methods §4.

### Colocalization

For each cis region we ran coloc.abf [43] over its five hypotheses; PP.H4 indexes a shared causal variant (colocalization) and PP.H1 an exposure-only signal. Priors, the cis window, and the single-causal-variant caveat (flagged at allelically heterogeneous loci such as LPA) are in Supplementary Methods §3.

### Reprogramming reversibility

Reprogramming datasets were scored with the chronological (Horvath, Hannum), phenotype (PhenoAge) and causality-enriched damage (DamAge/AdaptAge/CausAge, intercept-free) clocks; reversal was the reprogrammed − control difference, paired where an internal control existed. Processed betas were used where available, otherwise raw IDATs were noob-normalized (methylprep [44]); processing was validated against the authors’ own DNAmAge values, taken from the GEO sample characteristics (GSE142439, r=0.970 across all 16 samples). Datasets, internal controls, and the in-vivo mouse age-adjustment are in Supplementary Methods §5.

### Cross-species anchor and statistics

The Gompertz baseline prior was anchored by the cross-species mortality-rate-doubling-time–lifespan relation in AnAge [13] (`s1_crossspecies_tau.py`; Supplementary Methods §6). Analyses used Python 3.12 (numpy, pandas, scipy, statsmodels) with isolated environments for PyMC/arviz/emcee and methylprep, and R 4.3 for mammalian-array reading. Uncertainty is reported as 95% highest-density intervals for the Bayesian generator-matrix posteriors (NHANES and the discretized respecification), an equal-tailed 95% percentile interval for the HRS replication and for the prior-sensitivity sweep, percentile bootstrap 95% intervals for the subgroup and parameter-recovery analyses, and 95% confidence intervals for the frequentist analyses (Mendelian randomization, the reprogramming donor-level t-tests). Bayesian point estimates are posterior medians. OpenGWAS and Synapse access tokens are read from local files and are never hard-coded.

### Data and code

All primary data are public or provider-restricted (NHANES + Linked Mortality; HRS [restricted; aggregate-only, local]; GEO; UKB-PPP via Synapse syn51365303; deCODE; GWAS Catalog GCST006697; AnAge). Full numerical results are provided as the Supplementary Tables 1–18 and code is archived at Zenodo (DOI: 10.5281/zenodo.20790403). HRS analyses are aggregate-only per data-use terms; restricted HRS individual-level data and licensed pQTL files are not redistributed.

## Abbreviations

MR, Mendelian randomization; cis-pQTL, cis protein quantitative trait locus; coloc, colocalization; PP.H4, posterior probability of a shared causal variant; PP.H1, posterior probability of an exposure-only signal; GWAS, genome-wide association study; DNAm, DNA methylation; HDI, highest-density interval; MDE, minimum detectable effect; HR, hazard ratio; IIS, insulin/IGF-1 signalling; NHANES, National Health and Nutrition Examination Survey; HRS, Health and Retirement Study; UKB-PPP, UK Biobank Pharma Proteomics Project.

## Author Contributions

M.T. conceived the study, developed the generator-matrix and evaluation framework, performed the analyses, and wrote the manuscript. T.I. co-developed the reliability-theory / scaling framework and methodology and critically revised the manuscript. Both authors approved the submission.

## Supporting information

STROBE-MR checklist

## Acknowledgments

The authors used AI assistants in preparing this work: M.T. used Anthropic Claude for code development, analysis support, and language editing, and T.I. used OpenAI Chat-GPT 5.6 Sol (Codex) for an independent consistency and code review of the manuscript and analysis code. All data, analyses, figures, and text were verified by the authors, who take full responsibility for the content; no AI system was used to generate data or images.

We acknowledge the data providers. **HRS:** “The Health and Retirement Study (HRS) is sponsored by the National Institute on Aging (grant number NIA U01AG009740) and is conducted by the University of Michigan”; HRS Venous Blood Study, DNA-methylation, and RAND-linked mortality data were analysed locally in aggregate form under the HRS data-use terms (no individual-level data shared; cell sizes ≥ 5). **NHANES:** the National Center for Health Statistics (NCHS), Centers for Disease Control and Prevention, for the public NHANES data and Linked Mortality Files. **UKB-PPP:** only the publicly released plasma proteomic pGWAS summary statistics (Sun et al. 2023) were used, via Synapse (syn51365303); no individual-level UK Biobank data were accessed. **deCODE:** deCODE genetics/Amgen for the publicly released plasma-proteome pQTL summary statistics (Ferkingstad et al. 2021). **GWAS Catalog:** parental-lifespan summary statistics from the NHGRI-EBI GWAS Catalog (GCST006697; Pilling et al. 2017). **GEO:** DNA-methylation datasets (GSE165178, GSE165179, GSE142439, GSE247179, GSE190665); we thank the original data generators (cited). **OpenGWAS / Ensembl / AnAge:** outcome associations via the MRC IEU OpenGWAS database; variant annotation with Ensembl; cross-species ageing-rate data from the Human Ageing Genomic Resources (AnAge; Tacutu et al. 2018).

## Ethical Statement

This study analysed previously published public data (NHANES with Linked Mortality; GEO methylation; GWAS/pQTL summary statistics) and provider-restricted, de-identified individual-level data (HRS) under a data-use agreement; no new experiments on human participants or animals were performed. The source cohorts (NHANES, HRS) were conducted under their own institutional review board approvals and obtained informed consent from participants in the original studies. The present secondary analysis used only de-identified data: HRS individual-level data were analysed locally in aggregate form (cell sizes ≥ 5) under the HRS data-use terms, and no individual-level data were redistributed. The study was reviewed by the Research Ethics Committee of the Faculty of Medicine, Oita University, which determined on 3 July 2026 that it was exempt from formal ethical review.

## Consent

Not applicable to the present secondary analysis: informed consent was obtained from participants by the original cohort studies (NHANES, HRS), no identifiable individual-level data were used, and no additional consent was required for this de-identified secondary analysis.

## Data Availability

All primary data are public or provider-restricted (NHANES + Linked Mortality; HRS [restricted; analysed locally, aggregate-only]; GEO [GSE165178, GSE165179, GSE142439, GSE247179, GSE190665]; UKB-PPP via Synapse syn51365303; deCODE; GWAS Catalog GCST006697; AnAge). Analysis and figure code and all derived result tables are archived at Zenodo (DOI: 10.5281/zenodo.20790403) and provided as Supplementary Materials; restricted HRS individual-level data and licensed pQTL files (UKB-PPP, deCODE) are not redistributed and are obtained from their providers as described.

