## Supplementary material for "A generator-matrix model quantifies the limited contribution of measured biomarkers to human mortality acceleration": STROBE-MR checklist

§8. Supplementary Results (analyses subordinate to the three conditions) . . 10

**Supplementary Materials — Figure and Table Legends** **11**

### Supplementary Methods and Derivations

*Supporting information for:* "A generator-matrix model quantifies the limited contribution of measured biomarkers to human mortality acceleration."

This appendix contains the full mathematical formulation, derivations, priors, and sensitivity analyses. The main-text Results and Materials and Methods give the plain-language design; equation-level detail lives here and is cross-referenced as "Supplementary Methods §N."

---

#### §1. Generator-matrix / phase-type model of mortality acceleration

##### 1.1 State space and generator

We represent the organism as occupying hallmark-load states $s = (h_1, \dots, h_m)$ with $h_j \in \{0, \dots, L\}$ the discretized load on axis $j$, plus an absorbing death state $\partial$. The nine PhenoAge NHANES biomarkers were grouped, without collapsing to one dimension, into $m = 4$ hallmark axes: A1 inflammation/immune (CRP, white-cell count, lymphocyte %), A2 metabolic/nutrient-sensing (fasting glucose, albumin), A3 organ reserve (creatinine, alkaline phosphatase), A4 haematopoietic (mean cell volume, red-cell distribution width). State transitions follow a continuous-time Markov generator $Q$ with damage-progression rates $\lambda_j(\ell; \mathbf{x})$, repair rates $\mu_j(\ell; \mathbf{x})$, and an absorbing death rate $\kappa(s; \mathbf{x})$. Writing the transient block $T$ and exit vector $\mathbf{t}^0 = \kappa$, the age at death of an adult started at $a_0$ from initial distribution $\alpha$ is **phase-type**:

$$S(t) = \alpha\, e^{Tt}\mathbf{1}, \qquad f(t) = \alpha\, e^{Tt}\,\mathbf{t}^0, \qquad \mu(t) = f(t)/S(t).$$

Ergodicity makes the population distribution of age at death informative for $T$ without following individuals longitudinally; the role of unobserved frailty heterogeneity in population mortality is classical [34]. Phase-type and inhomogeneous-Markov representations of human mortality, including the interpretation of states as stages of biological aging, are well established [15,16]; those models are fit to mortality data alone, where the intensity matrix is only weakly identifiable, and regressing individual transition rates onto aging markers was noted there as a desirable but overparameterized extension. The present contribution is to break that identifiability with a joint likelihood that ties the stages to measured biomarker axes, and to use the fitted model to decompose Gompertz acceleration into biomarker-driven *visible* and *latent* components. The mortality hazard is adjusted log-linearly for covariates $\mathbf{x}$ — sex, BMI (standardized), and smoking status (ever/never), entered as additive log-hazard terms $b_{\text{male}}, b_{\text{BMI}}, b_{\text{smoke}} \sim \mathcal{N}(0, 1)$; never race or nationality (`bayes_unified_generator.py`). Smoking pack-years were not available in the pooled analytic extract, so binary ever/never smoking is used; the NHANES survey design (weights, strata, PSU) is addressed by a design-weighted Gompertz benchmark (§7) rather than by weighting the Bayesian model. For HRS, sex is included as a covariate; BMI and smoking were not available in the HRS analytic extract.

#### 1.2 Joint estimation and the visible/latent decomposition

For estimation and the visible/latent decomposition we used the joint formulation implemented in `bayes_unified_generator.py`: a per-axis drift $\lambda_j$ shared between (i) a state-dynamics likelihood, in which the cross-sectional age gradient of each standardized axis $z_{ij} \sim \mathcal{N}(b_j + \lambda_j(a_i - a_c), \sigma_j)$ identifies $\lambda_j$, and (ii) a proportional-hazard Gompertz mortality likelihood [35] in which accumulating damage raises the hazard, so that the same $\lambda_j$ contributes a **visible** acceleration $\sum_j \beta_j \lambda_j$ on top of a **latent** baseline $\gamma_{\text{base}}$. The total Gompertz acceleration is

$$\gamma = \gamma_{\text{base}} + \sum_j \beta_j \lambda_j, \qquad \text{latent fraction} = \frac{\gamma_{\text{base}}}{\gamma} = \frac{\gamma_{\text{base}}}{\gamma_{\text{base}} + \sum_j \beta_j \lambda_j}.$$

Priors: $\beta_j \sim \mathcal{N}(0, 0.5)$, $\lambda_j \sim \mathcal{N}(0, 0.03)$, $\sigma_j \sim \text{HalfNormal}(1)$, $\log \lambda_0 \sim \mathcal{N}(\log 10^{-3}, 2)$, and a weakly-informative prior $\gamma_{\text{base}} \sim \mathcal{N}(0.08, 0.03)$ centred on the cross-species mortality-rate-doubling-time–lifespan relationship recorded in the public AnAge database [13] (§6). Posterior sampling used PyMC [36] NUTS (4 chains, 2000 tune, 1000 draws, target-accept 0.99, `adapt_diag` initialization); convergence was assessed by $\hat{R}$ (all = 1.00) and effective sample size (all bulk-ESS > 3000,

0 divergences). For the HRS replication (`hrs_bayes_generator.py`), whose DNA-methylation clock axes drift on a larger per-year scale than the standardized blood axes, the drift and baseline-rate priors were widened accordingly — $\lambda_j \sim \mathcal{N}(0, 0.3)$ and $\log \lambda_0 \sim \mathcal{N}(\log 0.02, 2)$ — while $\beta_j$, $\sigma_j$, $\gamma_{\text{base}}$, the sex covariate and the sampler settings were identical to NHANES. Adding the sex and BMI covariates required `adapt_diag` initialization and the higher target-accept for stable sampling (with default jitter initialization the more strongly correlated posterior diverged), but this is a sampler-geometry adjustment, not a change to the model or priors; the frequentist point estimate is identical and robust (§7). As a check robust to the non-differentiable matrix-exponential likelihood, the full phase-type first-passage model was also fitted by ensemble MCMC (emcee [37]; on the efficiency of non-local trial moves in Markov chain Monte Carlo, see [38]). The mean-field limit $E[\text{death age}] = \alpha(-T)^{-1}\mathbf{1}$ recovers a disposable-soma / reliability decomposition $\tau_{\text{eff}} \approx \tau_0\, g_{\text{maintenance}}\, g_{\text{allocation}}$ [12,39], of which the present model is the dynamical, distribution-level generalization.

#### 1.3 Two estimators and the differential test (honest scope of "generator-matrix")

The headline latent fraction is the estimate of the **joint biomarker-and-mortality model** of §1.2 (a Gompertz proportional-hazard mortality coupled to the cross-sectional biomarker-drift likelihood): 92.7% [92.2–93.2] (NHANES), 91.5% [89.7–93.3] (HRS). As a sensitivity re-specification, the same decomposition was recomputed from **discretized hallmark loads with a mean-field visible term** (`phasetype_decomposition_bayes.py`), matched to the primary complete-case sample ($n = 23{,}512$) and to the same sex/BMI/smoking adjustment: latent 89.1% [88.0–90.0] at $L = 3$ (88.7–90.0% across $L \in \{2, 3, 4\}$), a 3.6-percentage-point difference from the primary. **Honest scope:** this is *not* a person-level phase-type first-passage likelihood. It discretizes each axis into $L + 1$ quantile levels, obtains $\lambda_j$ from a load-on-age regression and $(\gamma_{\text{base}}, \beta_j)$ from a Poisson survival GLM (Laplace normal approximation), and forms $\gamma_{\text{base}}/(\gamma_{\text{base}} + \beta \cdot \lambda)$ over draws; the $(L+1)^4$-state age propagation is a point-estimate consistency check only (propagated $d \log h/da = 0.079$ vs $\gamma_{\text{base}}$+visible$= 0.085$, the difference being the survivorship term), and the repair transitions drawn in Fig. 1 are not part of this script. It is therefore a differently-specified sensitivity analysis, not an independent implementation of the same estimand. We report the individual estimates (92.7% primary NHANES; 91.5% HRS; 89.1% discretized mean-field) rather than a single pooled range. The full person-level phase-type likelihood (`phasetype_person_likelihood.py`) was not computationally tractable at this state-space size with gradient-free optimizers (a diagnostic confirmed the model is identifiable, `phasetype_identifiability_diag.py`); the primary estimate is therefore the joint proportional-hazard model, cross-checked against the phase-type formulation. The engine oracle (`phasetype_oracle_test.py`) passed 14/14 (Erlang $k = 1, 3, 5$ to machine precision; constant-death vs ODE $5.5\times10^{-7}$; invariants on 30 random generators).

#### 1.4 Structural verification (Lean)

The decomposition's structural invariants were machine-checked in the Lean 4 proof assistant [46] with Mathlib [47] (`verification/lean/ShareDecomposition.lean`, with the project-to-lemma mapping in `verification/lean_verification.md`; both are included in the Zenodo deposit). The development is `sorry`-free and introduces no axioms of its own (7 theorems across the namespaces `LeanVerify.Share` and `LeanVerify.Detection`), and was checked with Lean 4 v4.32.2 and Mathlib pinned at rev v4.32.2, as recorded in the deposited `lean-toolchain` and `lakefile.toml`. With share$(a, b) = a/(a + b)$ ($a = \gamma_{\text{base}}$, $b = \sum_j \beta_j \lambda_j$): the latent fraction lies in $[0, 1]$ (`share_nonneg`, `share_le_one`); it $\to 1$ as the visible part $\to 0$ (`share_eq_one_of_b_zero`, `share_tendsto_one_of_b_to_zero`), $= 0$ when $\gamma_{\text{base}} = 0$ (`share_eq_zero_of_a_zero`), and is monotonically decreasing in the visible part (`share_antitone_in_b`). These formalize the numerical property tests and confirm the decomposition is structurally sound (sign and limits as expected); the *value* of the fraction is a data/estimator question addressed by §1.2–1.3, not by the formal proof.

---

### §2. Mendelian randomization

Two-sample cis-pQTL Mendelian randomization [41] used genome-wide-significant ($p < 5 \times 10^{-8}$) cis variants from UKB-PPP (REGENIE summary statistics, GRCh38; effect allele = ALLELE1; cis window ±1 Mb of the gene) and deCODE (SomaScan, GRCh38, with rsIDs; cis lead searched within ±200 kb, `mr_coloc_decode.py`). For UKB-PPP the cis lead SNP was mapped to an rsID via the Ensembl GRCh38 REST API; outcome effects came from the parental-lifespan GWAS [22] (by rsID through the MRC IEU OpenGWAS API [42], or by GRCh38 position from the harmonised GWAS-Catalog file). Effects were harmonized to the protein-increasing allele by allele matching and strand complementation (`mr_ukbppp.py`); effect-allele frequencies were not used, and the five UKB-PPP lead variants reported here are non-palindromic. The Wald ratio was computed with a delta-method standard error and is expressed on the outcome's native scale, as **SD of the rank-normalized Martingale residual of combined parental attained age per SD higher protein** (abbreviated SD/SD); **the outcome is not calibrated in years**, so no year-equivalent is reported. The primary estimates reported in the main text are single-lead Wald ratios. Where two or more independent cis instruments existed (e.g. GDF15, three instruments), inverse-variance-weighted MR with MR-Egger intercept and weighted-median sensitivity analyses were additionally run (`mr_latent_causation.py`, `MR_latent_causation.csv`); these did not change the null conclusions.

**Sample overlap.** The UKB-PPP exposure sample (n=54,219) is contained within the UK Biobank outcome sample (n=389,166), so the two-sample independence assumption is violated. Two points bound the consequence. First, with overlapping samples the bias acts toward the *confounded observational* association rather than toward zero; because these markers are strongly associated with mortality observationally, overlap would move estimates away from the null, so the observed nulls are the conservative result. Second, the instruments are strong (F = 1499–20031), making weak-instrument bias negligible. The deCODE (Icelandic) exposure arm shares no participants with the outcome and reproduces the same nulls; note, however, that both platforms share the *same* outcome GWAS, so they provide exposure-platform — not outcome — replication. Exposures were the latent's measurable components (GDF15, cystatin C/CST3, PAI-1/SERPINE1, TIMP1, B2M, IL6) and the growth-signalling/IIS axis (IGFBP1/2/3, GH1, GHR, IGF1R; IGF-1 from deCODE). Known-causal positive controls (LPA, IL6R) were analysed identically to calibrate power, and instrument strength was summarized by the F-statistic.

**Outcome orientation.** The continuous parental-lifespan outcome (`ebi-a-GCST006697`) is provided on the Martingale-residual scale, on which a *positive* native effect is *detrimental* (shorter parental life): the ApoE ε4 allele, which shortens life, carries a positive native effect on this scale in this dataset (APOE rs429358). We therefore re-oriented all continuous-outcome Wald estimates so that a positive effect *lengthens* life (`mr_orient_lifespan.py`), consistent with the binary longevity outcomes (`ebi-a-GCST006698`, `ebi-a-GCST003395`, natively oriented positive = more longevity), and verified as an oracle that the known-causal positive controls then carry biologically correct signs (LPA harmful, β<0; IL6R protective, β>0). This re-orientation changes only the sign of point estimates and their confidence intervals; magnitudes, standard errors, p-values, colocalization posteriors, the null conclusions, and the minimum-detectable-effect power analysis (which use the effect magnitude) are unchanged.

**Directionality (MR-Steiger).** To assess directionality (whether lifespan could influence protein rather than the reverse), we applied the MR-Steiger test (Hemani, Tilling & Davey Smith, PLoS Genet 2017): for each instrument we compared the variance it explains in the exposure, $R^2_{\text{exp}} = z^2_{\text{exp}}/(z^2_{\text{exp}} + N_{\text{exp}})$, and in the outcome, $R^2_{\text{out}} = z^2_{\text{out}}/(z^2_{\text{out}}+N_{\text{out}})$ ($N_{\text{exp}} = 54{,}219$ UKB-PPP; $N_{\text{out}} = 389{,}166$ Pilling 2017), with a Steiger $z$ comparing the two correlations. Because the exposure sample lies within the outcome sample, the independent-samples assumption behind the Steiger $z$ is violated and its $p$-value is approximate; the robust quantity is the $R^2$ ratio (`mr_steiger.py`). For each of the five UKB-PPP instruments tested (GDF15, CST3, SERPINE1, IL6R, LPA), $R^2_{\text{exp}} \gg R^2_{\text{out}}$ (ratio $2.2 \times 10^2$–$2.5 \times 10^5$; all Steiger $p < 10^{-27}$), supporting the protein-to-lifespan direction rather than the reverse.

**Multiple testing.** Seven non-control aging-marker exposures were tested on the primary platform (UKB-PPP × parental lifespan): GDF15, CST3, SERPINE1, IGFBP1, IGFBP3, GHR, IGF1R. This family was fixed in advance and is not enlarged by the deCODE analyses, since enlarging a family weakens the correction. Two exposures reached nominal significance — GHR ($p = 0.018$) and SERPINE1 ($p = 0.022$) — and **neither survives Bonferroni correction** ($\alpha = 0.05/7 = 0.007$) **nor Benjamini-Hochberg control of the false-discovery rate at 5%** (smallest $q = 0.076$; `mr_multiplicity.py`, S-Table 18). Reporting both matters because Bonferroni alone invites the objection that it is over-conservative: here the permissive correction rejects nothing either. We do not invoke colocalization for these two signals; as §3 sets out, colocalization has no demonstrated sensitivity in this design, and for SERPINE1 the distinct-causal-variant posterior never exceeds 0.082, so a linkage-disequilibrium explanation is not supported. No non-control marker shows a multiplicity-robust signal. By contrast, the two positive controls survive any reasonable correction (LPA $p = 9 \times 10^{-12}$; IL6R $p = 2.8 \times 10^{-5}$), confirming the design detects genuine causal effects at these instrument strengths. The conclusion — that the measurable components are consistent with a downstream-marker interpretation — is thus not an artefact of examining multiple markers: it holds under, and is strengthened by, correction for multiplicity.

---

### §3. Colocalization

For each cis region we ran coloc.abf [43], comparing the pQTL and lifespan GWAS over five hypotheses (H0 no association; H1/H2 one trait only; H3 distinct causal variants; H4 a shared causal variant). Per-SNP approximate Bayes factors used $z = \beta/\text{se}$ and $V = \text{se}^2$ with prior variance $W = 0.15^2$ and priors $p_1 = p_2 = 10^{-4}$, $p_{12} = 10^{-5}$, over the ±100 kb cis window, matched to the lifespan GWAS by GRCh38 position (computed in log-space with the log-sum-exp identity). PP.H4 indexes colocalization and PP.H1 an exposure-only signal. All five posteriors are reported (S-Table, coloc).

**Colocalization is descriptive here, not corroborative.** Its sensitivity is not established in this design, for three reasons. (i) *Neither positive control colocalized.* At LPA, PP.H3 = 1.000 and PP.H4 = 0.000, the expected failure mode of the single-causal-variant assumption at an allelically heterogeneous locus (KIV-2 copy-number and multiple independent cis signals); at IL6R, PP.H4 = 0.345. A method that does not colocalize two proteins with established causal effects on the outcome cannot be used to argue that a low PP.H4 indicates the absence of a causal effect. (ii) *The verdict depends on an analyst-set prior.* We repeated coloc.abf over four cis-windows (±50, 100, 200, 500 kb) and five values of $p_{12}$ $(10^{-6}, 5 \times 10^{-6}, 10^{-5}, 5 \times 10^{-5}, 10^{-4})$, holding $p_1 = p_2 = 10^{-4}$ and $W = 0.15^2$ (`coloc_sensitivity.py`; S-Table 17). Because per-SNP approximate Bayes factors are prior-independent, the 100 cells were recomputed from a single extraction at the widest window; the published cell (100 kb, $p_{12} = 10^{-5}$) reproduces the main analysis to within 0.01 (an oracle assertion in the script). Whether IL6R colocalizes is entirely set by $p_{12}$ (PP.H4 = 0.047, 0.20, 0.35, 0.72, 0.84 from $10^{-6}$ to $10^{-4}$), whereas the window changes PP.H4 by ≤0.005. (iii) *Low PP.H4 here means low outcome-side power, not distinct variants.* For CST3 and SERPINE1 the posterior mass sits on H1 (exposure only: 0.785–0.988 and 0.738–0.989 across the grid), i.e. the lifespan GWAS carries no detectable signal in the region. The distinct-causal-variant hypothesis is therefore *not* supported for SERPINE1 (PP.H3 ≤ 0.082 everywhere), so its nominal MR association cannot be attributed to linkage disequilibrium, as an earlier version of this work stated. GDF15 is the only marker locus at which H3 is robustly favoured (0.589–0.660). We therefore report colocalization as a descriptive companion to MR and rest no claim on it.

---

### §4. Null informativeness: minimum detectable effect (power)

For a Wald-ratio MR estimate with standard error SE, the two-sided test at level $\alpha$ rejects when $|\hat{\beta}| > z_{1-\alpha/2}\,\text{SE}$; under a true effect $\beta$ the power is $\Phi(\beta/\text{SE} - z_{1-\alpha/2}) + \Phi(-\beta/\text{SE} - z_{1-\alpha/2})$. The minimum detectable effect (MDE) at power $1-\eta$ is therefore

$$\text{MDE} = (z_{1-\alpha/2} + z_{1-\eta})\,\text{SE},$$

with $\alpha = 0.05$ (two-sided) and power $0.80$ giving $\text{MDE} = (1.959964 + 0.841621)\,\text{SE} = 2.801585\,\text{SE}$ (`mr_null_power.py`). A null with MDE $\leq$ a positive control's effect means an effect the size of a *known* causal protein would have been detected. For the strongly-instrumented markers the MDE was 0.010–0.023 SD/SD, at or below the LPA effect (0.027 SD/SD): GDF15, CST3, IGFBP3 and GHR each had ≥80% power against an LPA-scale effect. The weakly-instrumented markers (SERPINE1, IGFBP1, IGF1R) had MDE above the positive-control effects, so this design does not resolve them; we do not claim to exclude effects as small as the smaller IL6R effect (0.009 SD/SD). Per-protein MDEs are in S-Table (power). The derivation core, that the MDE solves the power equation exactly, was machine-checked in Lean (`DetectionThreshold.mde_solves_power`; see §1.4 and `notes/lean_verification.md`).

---

### §5. Reprogramming reversibility (clock battery)

Processed EPIC/450K beta matrices were used where available; raw IDATs were processed to noob-normalized betas with methylprep [44] in an isolated Python 3.10 environment. Each dataset was scored with the chronological (Horvath, Hannum), phenotype (PhenoAge) and causality-enriched damage (DamAge/AdaptAge/CausAge, intercept-free, so only group differences are interpreted) clocks. Reversal was the reprogrammed − control difference, paired where an internal control existed (GSE165178: SSEA4$^{+}$ reprogramming vs CD13$^{+}$ non-reprogrammed cells; GSE142439: Treated vs Normal per donor). For the in-vivo mouse data (GSE190665, mammalian array, clock1 [40]) the treatment effect was age-adjusted by regression because treated animals were chronologically older. Processing was validated against authors' reported DNAmAge (GSE142439 r = 0.97). Epigenetic clocks were scored with a streaming pure-Python engine (`clocks.py`) implementing Horvath (with its anti-transform), Hannum, and PhenoAge, and, for reprogramming, the Ying et al. [18] DamAge/AdaptAge/CausAge causal clocks; mouse arrays used the universal mammalian clock [40]. EPICv2 probes were collapsed to base cg identifiers; clock CpGs missing from a matrix were mean-imputed from a reference panel (`--impute-ref`) and per-clock coverage reported.

**Donor-level inference (corrected 2026-09-17).** Both Gill datasets sample the *same* donors at several days (GSE165178: 4 donors × days 9/11/15, 11 matched SSEA4/CD13 pairs; GSE165179: 3 donors × days 10/13/15/17 × 2 experiments, 13 matched reprogrammed/negative-control pairs), so the matched pairs are nested within donor and are not independent. An earlier analysis treated them as independent (a paired $t$-test over the 11 pairs) and, for GSE165179, compared unpaired group means that ignored donor, day and experiment. We therefore re-analysed both datasets at the donor level (`rev_donor_level.py`, `rev_donor_level_mptr.py`): the reprogrammed − control difference is formed within each donor (and day/experiment), averaged over days within donor, and tested with a one-sample $t$-test on the donor means; we also report the difference stratified by day. Results are in `REV_GSE165178_donorlevel.csv` and `REV_GSE165179_donorlevel.csv` and are the values quoted in the main text and Table 3. Two consequences are worth stating plainly. First, in MPTR the dissociation is day-stable: DamAge lies in the opposite direction to Horvath at every timepoint (+13.3 to +17.8 across days 10–17). Second, in the Sendai series it is day-dependent: during the partial, identity-retaining window (days 9–11) neither clock is resolved with four donors, whereas at day 15 — as full iPSC identity is approached — both the chronological clock (−47.2, $p = 0.001$) and the damage clock (−24.4, $p = 0.030$) reverse. With three to four donors per protocol these tests have low power, and a non-significant donor-level result is **not** evidence of equivalence.

**DNAm surrogate localization of the latent (HRS)**

In HRS, protein-trained DNAm surrogate modules (inflammaging = DNAm CRP+GDF15+B2M; senescence = DNAm PAI-1+TIMP1; metabolic = DNAm HbA1c+leptin; renal = DNAm cystatin C) were added one at a time to the four blood axes (five-axis Bayesian generator models) and the reduction in latent fraction recorded; epigenetic ages, DunedinPACE and GrimAge were additionally Cox-modelled for mortality beyond age and blood axes. Whole-blood RNA-seq inflammaging/senescence gene modules (e.g. CDKN2A, CDKN1A, SERPINE1, GDF15, IL6, CXCL8, TNF, NFKB1) were tested in parallel by Cox regression.

---

### §6. Cross-species anchor and statistics

The Gompertz baseline prior was anchored by the cross-species mortality-rate-doubling-time–lifespan relationship in the public AnAge database [13] (`s1_crossspecies_tau.py`). Analyses used Python 3.12 (numpy, pandas, scipy, statsmodels), with isolated environments for PyMC/arviz/emcee (numpy < 2) and methylprep (Python 3.10, numpy < 2, pandas < 2), and base R 4.3 for `.RDS`/mammalian-array reading. Uncertainty is reported as 95% highest-density intervals (generator-matrix posteriors), equal-tailed 95% percentile intervals (HRS replication, prior sweep), percentile bootstrap intervals (subgroup, parameter recovery), or 95% confidence intervals (frequentist). OpenGWAS and Synapse access tokens are read from local files and are never hard-coded.

---

### §7. Sensitivity analyses

**Prior sensitivity (latent fraction).** On the covariate-adjusted primary model (n = 23,512), the latent fraction is robust to the $\gamma_{\text{base}}$ prior centre: shifting the prior mean across $\mathcal{N}(0.04, 0.03)$, $\mathcal{N}(0.08, 0.03)$, $\mathcal{N}(0.12, 0.03)$ all returned 92.7% [92.2–93.2] with $\gamma_{\text{base}} \approx 0.088$, $\hat{R} = 1.00$, $\text{ESS}_{\text{bulk}} \sim 6400\text{–}7100$, $\text{ESS}_{\text{tail}} \sim 3300\text{–}3450$, 0 divergences (`prior_sensitivity_covariate.py`; Supplementary Table 15). Very diffuse priors on $\gamma_{\text{base}}$ ($\mathcal{N}$ with $\sigma = 0.30$, or HalfNormal) weaken convergence — consistent with the sampler-geometry note in §1.2 — and their point estimates are not interpretable; they are reported as a computational note, not as evidence.

**Identifiability and interval calibration.** Because inferring mechanism from mortality alone is many-to-one, we tested directly whether the latent fraction is *identified* by the joint biomarker-and-mortality likelihood. Simulating datasets from the generative model at a grid of known latent fractions (60–95%) and recovering each with the estimator, recovery was unbiased and monotonic (|bias| $\leq 1.3$ pt across the grid) and the nonparametric bootstrap 95% CI covered the truth at a mean of 0.93 (0.88–0.98; the mild under-coverage occurs only at the lowest, highest-variance latent, as expected for the percentile bootstrap). The latent fraction is therefore identified by the joint likelihood, not an artefact of a flat likelihood ridge (`identifiability_coverage.py`; Supplementary Table 16). This is a frequentist calibration of the fast estimator (which agrees with the Bayesian model to $< 0.5$ pt, §1.3); a full Bayesian simulation-based calibration is left to future work.

**Discretization sensitivity (load levels $L$).** The phase-type latent fraction was stable across $L \in \{2, 3, 4\}$: 90.0% / 89.1% / 88.7% (`phasetype_decomposition_bayes.py`).

**Covariate adjustment.** Adjusting the mortality model for sex, BMI and smoking status left the latent fraction essentially unchanged (unadjusted 91.9% vs fully adjusted 92.2% by design-matched Poisson GLM; the Bayesian primary is reported in the main text).

**Survey-design benchmark.** The pooled NHANES analytic model is unweighted. Re-estimating the same proportional-hazard decomposition with MEC examination sampling weights (WTMEC2YR, pooled across cycles) via a design-weighted Poisson GLM changed the latent fraction by less than two percentage points (unweighted 91.9% vs survey-weighted 90.1%; `s5_survey_benchmark.py`), so the decomposition is robust to the complex survey design. (Strata/PSU affect variance, not this point estimate.)

**Two-cohort replication.** The decomposition replicated in an independent cohort and period (HRS, prospective mortality): 91.5% [89.7–93.3] vs 92.7% [92.2–93.2] in NHANES.

**Conditional expected-event-count diagnostic (in place of a calibrated posterior predictive check).** The mortality likelihood is the Poisson-process (Aalen) form $\delta_i \log h_i(u_1) - H_i$, so mortality is modelled as a counting process with integrated intensity $H_i = \int_{u_0}^{u_1} h_i$, and the internally-consistent predictive quantity is the expected event count $\mathbb{E}[N_i] = H_i$ (not the Bernoulli $1 - e^{-H_i}$, which saturates for the small fraction of persons whose integrated hazard exceeds 1 over long follow-up from the top-coded oldest ages). We replicated datasets from the same process — for each of 600 posterior draws, $y_{\text{rep}}(\text{band}) \sim \text{Poisson}(\sum_i H_i)$ — giving predictive intervals that include Poisson sampling and posterior uncertainty. The aggregate death count was reproduced (4,829 observed vs 4,827 expected). **This is weak evidence of calibration**: with a freely fitted baseline scale the score equations of this likelihood force the total expected count close to the observed total within the same sample, so aggregate agreement is close to automatic. Band by band, only 45–55, 55–65 and 75–85 fell inside the 95% replication interval; **three of six bands — 20–45, 65–75 and 85+ — fell outside it**, the largest discrepancy being 65–75 (1,322 observed vs 1,439.6 expected, interval [1,354, 1,525]). The misfit is therefore *not* confined to the extremes. NHANES age top-coding also differs by cycle (85 in 1999–2006, 80 in 2007–2010), so the oldest band mixes differently-censored ages. Finally, the replication draws band counts as Poisson$(\sum_i H_i)$ conditional on each person's observed follow-up, which is not the same generative process as a single-event survival model with administrative censoring; we therefore report this as a **conditional expected-event-count / residual diagnostic**, not a calibrated survival posterior predictive check, and leave a censoring-aware survival PPC to future work. It bears on absolute-count calibration rather than on the acceleration ratio that defines the latent fraction (Supplementary Figure 4, Supplementary Table 13; `bayes_unified_generator.py`, `fig_ppc.py`). Verification: a known-solution oracle confirms the expected-count PPC is unbiased on data generated from the same Poisson process (`gate_ppc_stratified.py`).

**Subgroup robustness.** The latent fraction was re-estimated within sex, age group ($< 65$ / $\geq 65$; each retaining enough age span for the Gompertz slope) and NHANES cycle using the fast frequentist Poisson-GLM estimator (the double-implementation test shows it agrees with the Bayesian model to $< 0.5$ pt), with a nonparametric bootstrap 95% CI (400 resamples) per stratum. Covariates were adjusted as in the primary model, dropping any covariate constant within a stratum. The latent fraction ranged 86.7–95.5% across all ten strata (overall 92.2%): female 95.5% [93.7–96.3] / male 86.7% [85.1–88.0]; age $< 65$ 89.7% [87.6–90.9] / $\geq 65$ 91.5% [88.7–93.4]; cycles 87.5–94.2%. The latent thus dominates the mortality acceleration in every subgroup and is not an artefact of any single sex, age band or survey wave (Supplementary Figure 5, Supplementary Table 14; `stratified_latent.py`). A subgroup-comparability oracle confirms the stratum estimator recovers the same latent across subgroups with differing covariate composition (`gate_ppc_stratified.py`).

---

### §8. Supplementary Results (analyses subordinate to the three conditions)

These two analyses extend, and are reported in support of, the three main-text conditions; they are placed here to keep the main text on the single account → cause → reverse argument.

**§8.1 DNA-methylation systemic-signature localization of the latent (supports the first, accounting condition).** To localize the latent, mechanism-mapped DNA-methylation surrogate modules (protein-trained, hence less circular than mortality-trained composite clocks) were added to the four blood axes in HRS, one at a time. The latent shrank most with a renal (DNAm cystatin C) module (−26.5 pt), then inflammaging (DNAm CRP+GDF15+B2M, −20) and senescence (DNAm PAI-1+TIMP1, −17), while a metabolic module did nothing (0). Generic chronological epigenetic age (Horvath) added nothing beyond blood (mortality HR 1.03, NS); a mortality-trained composite (GrimAge) captured ~2/3 of the latent but is intrinsically circular. Whole-blood transcription of the same inflammaging/senescence genes did not capture the latent (mortality HR ≈ 1.0). The latent therefore behaves as a slowly accumulating, DNAm-integrated systemic load (inflammaging/senescence/renal), not a transcriptional snapshot, and is invisible to generic chronological clocks (Supplementary Figure 3, Supplementary Tables 3–4; methods in §5).

**§8.2 The growth-signalling (insulin–IGF/hyperfunction) axis also shows no**

**detectable causal effect (extends the second, causation condition).** Because hyperfunction (B-1) locates the driver in nutrient/growth (mTOR–IIS) signalling, we tested the MR-tractable proxy of that axis, circulating IGF-axis and GH proteins. The IGF-1 carrier IGFBP3 carried a very strong cis-pQTL (lead $-\log_{10}p = 837$) and was null for lifespan with ≥80% power against an LPA-scale effect (Supplementary Table 12). GHR's nominal association (p=0.018) does not survive correction for the seven markers tested. IGFBP1 and IGF1R are underpowered (minimum detectable effects 0.14 and 0.06 SD/SD) and are not resolved by this design. Direct IGF-1 (deCODE) cannot be validly instrumented against this outcome GWAS: the lifespan GWAS has no coverage at its cis lead variant, and under the pre-specified fallback rule (strongest covered cis variant by exposure-side strength) all three covered genome-wide-significant cis variants prove to be rare (MAF 0.0037–0.0048). The Wald ratio they give ($\beta = +0.058$ [−0.017, +0.133], p = 0.13; MDE 0.107 SD/SD) is therefore reported to document that IGF-1 is untested, not null (`mr_igf1_fallback.py`; S-Table 18). The second aptamer (8406_17) has no genome-wide-significant cis variant at all. Thus no measurable circulating growth-signalling proxy shows a supported causal effect, and the one strongly instrumented proxy excludes an effect of positive-control magnitude, consistent with a downstream-marker rather than causal role for this axis in human longevity (Fig. 2; Table 2; Supplementary Table 7). Circulating pQTL-MR indexes the IIS axis but cannot measure intracellular mTOR activity; this does not refute the hyperfunction mechanism but shows that no detectable causal effect is carried by its measurable circulating proxy — the same latency in accessible measures seen for A and B-2.

### Supplementary Materials — Figure and Table Legends

*Companion to the manuscript "A generator-matrix model quantifies the limited contribution of measured biomarkers to human mortality acceleration." Supplementary Methods are provided separately (Supplementary_Methods). Supplementary Figures are provided as S1_Fig–S6_Fig (TIFF); Supplementary Tables as S1_Table–S16_Table (xlsx).*

*All Supplementary Materials files are derived or aggregate results computed in this study; no individual-level or restricted primary data are included. HRS-derived tables are aggregate (cell sizes ≥ 5) under the HRS data-use terms. Primary data are obtained from the sources cited under Data Availability.*

**Supplementary Figure 1. Generator-matrix decomposition of mortality acceleration.** 92.7% latent versus 7.5% visible (four measured axes with $\beta \cdot \lambda$ contributions). *Source:* NHANES 1999–2010 + Linked Mortality. (`fig_latent_decomp`; `S1_Fig`)

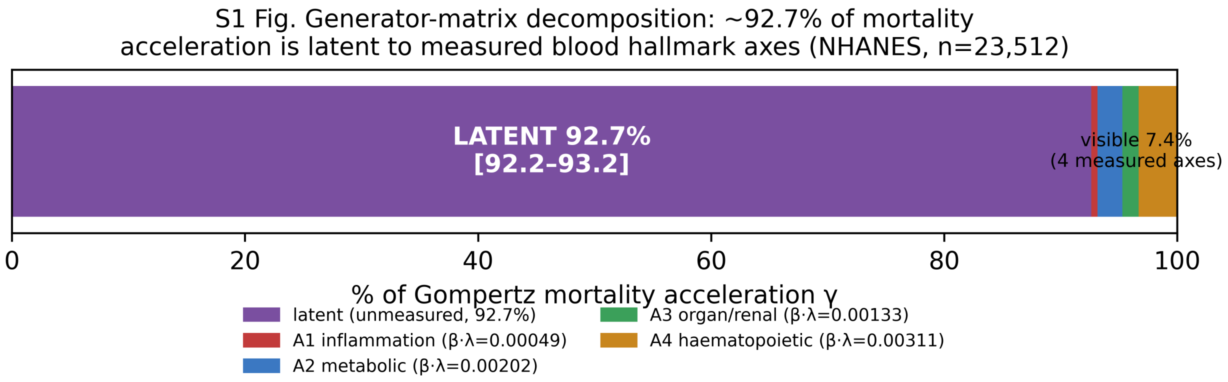

**Supplementary Figure 2. Two-cohort replication of the latent fraction.** NHANES 92.7% / HRS 91.5%. *Source:* NHANES + HRS (aggregate). (`fig_cohort_replication`; `S2_Fig`)

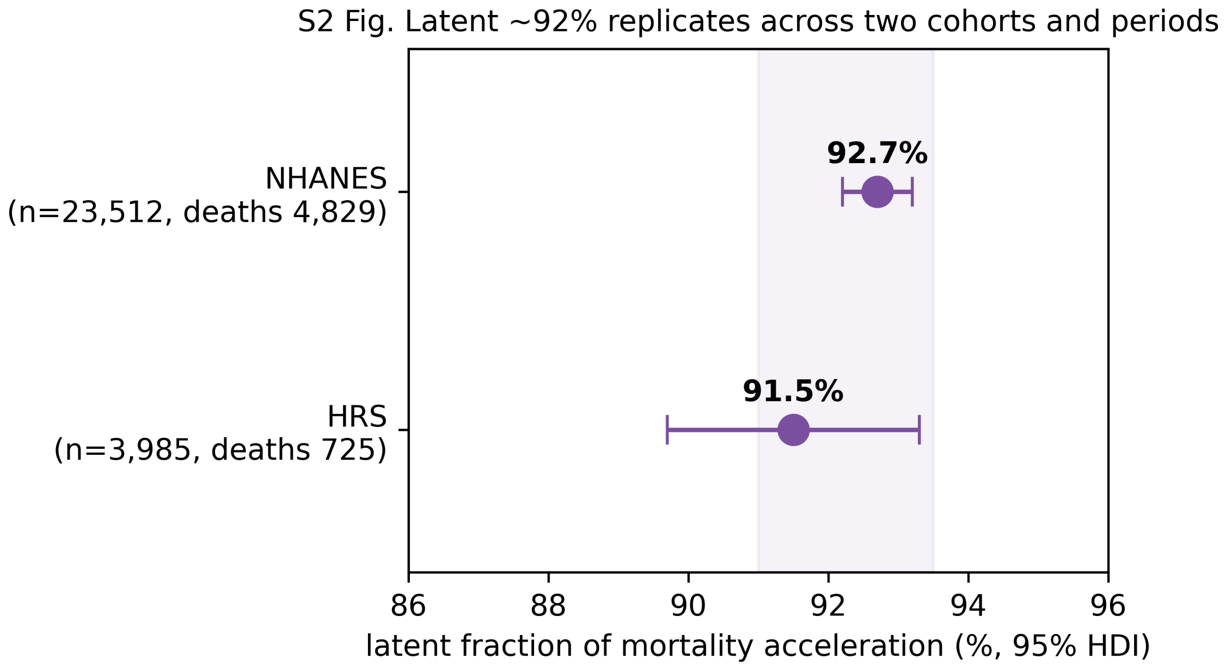

**Supplementary Figure 3. DNAm systemic-signature decomposition of the latent.** Renal −26.5 / inflammaging −19.8 / senescence −17.0 / metabolic 0 pt; whole-blood RNA-seq not significant. *Source:* HRS (aggregate). (`fig_dnam_modules`; `S3_Fig`)

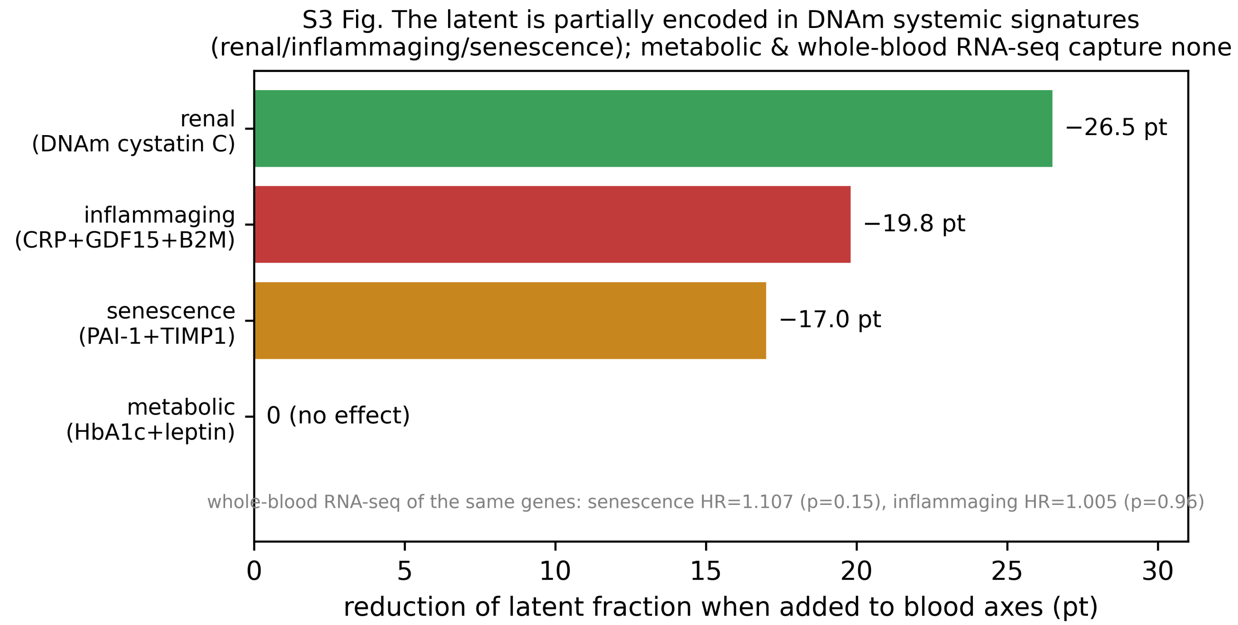

**Supplementary Figure 4. Posterior-predictive calibration of the primary NHANES model.** Observed versus posterior-predicted death counts by age band (95% predictive band, Poisson-process replication). Aggregate observed 4,829 versus expected 4,827 (close to automatic with a freely fitted baseline); three of six bands (20–45, 65–75, 85+) fall outside the 95% replication interval, so the misfit is not confined to the extremes. Reported as a conditional expected-event-count / residual diagnostic, not a calibrated survival posterior predictive check. *Source:* NHANES 1999–2010 + Linked Mortality. (`fig_ppc.py`; `ppc_calibration.csv`; `S4_Fig`)

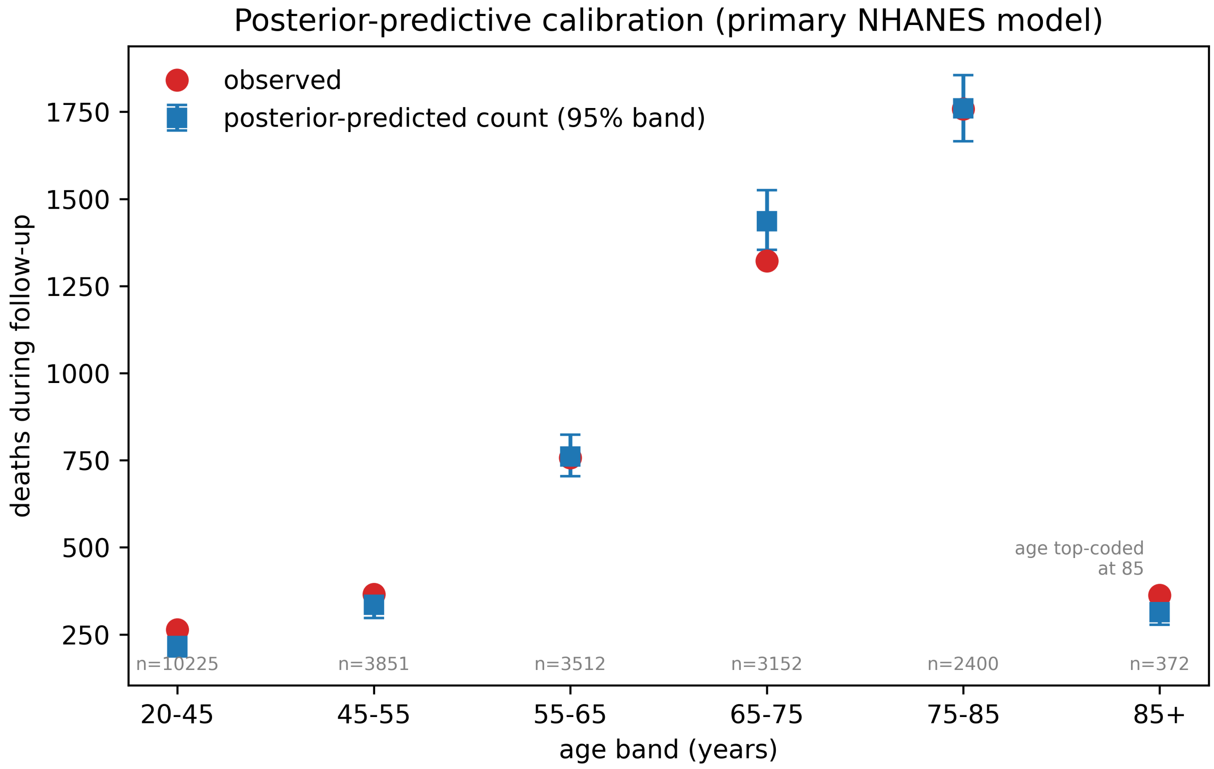

**Supplementary Figure 5. Subgroup robustness of the latent fraction.** Latent fraction (bootstrap 95% CI) re-estimated within sex, age group (<65 / ≥65) and NHANES cycle; range 86.7–95.5% (overall 92.2%). *Source:* NHANES 1999–2010 + Linked Mortality. (`fig_stratified.py`; `stratified_latent.csv`; `S5_Fig`)

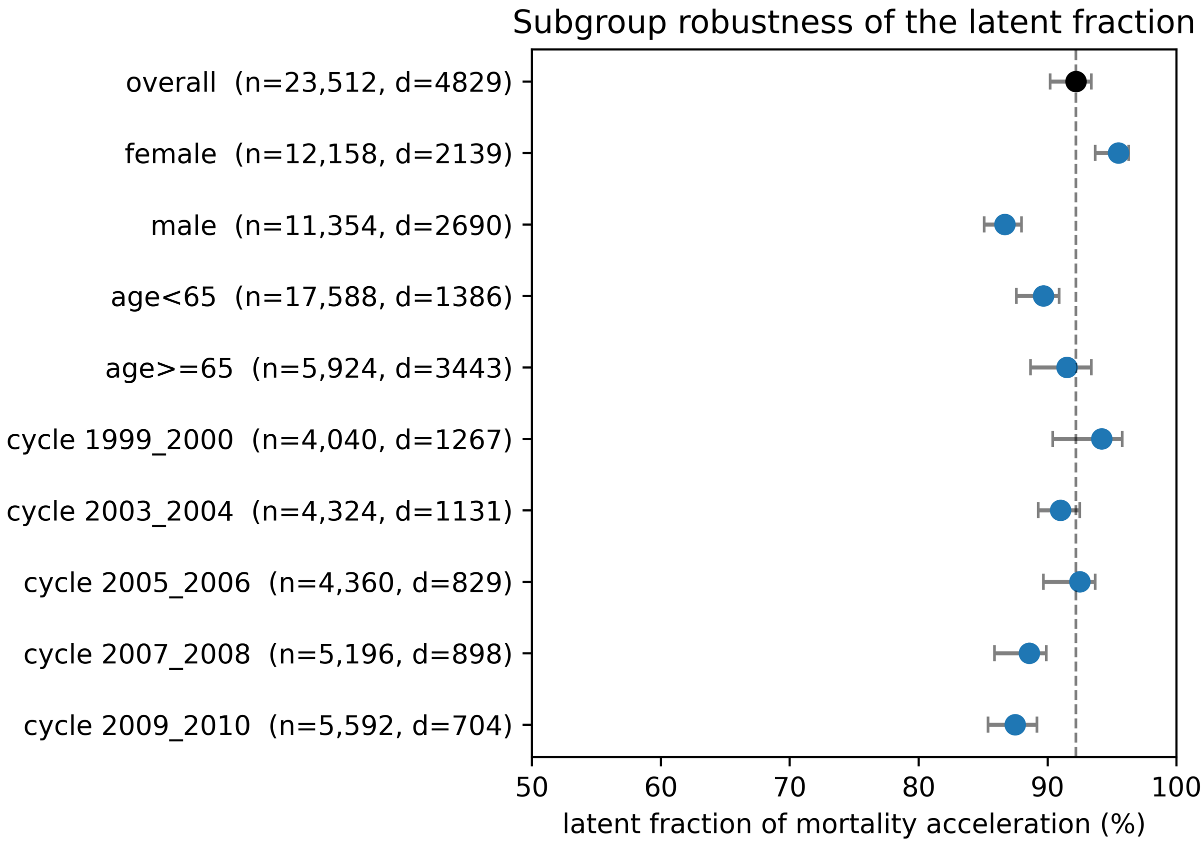

**Supplementary Figure 6. Primary model versus a discretized mean-field re-specification.** Latent fraction from the primary joint biomarker+mortality model (92.7%) versus a discretized hallmark-load / mean-field re-specification of the same decomposition, matched to the primary complete-case sample and covariates (L=3, 89.1%; L=2 and L=4 shown as discretization sensitivity); $|\Delta|$ = 3.6 pt. This is a sensitivity comparison, not an independent first-passage implementation. *Source:* NHANES 1999–2010 + Linked Mortality. (`fig_phasetype_correspondence.py`; `bayes_unified_summary.csv`, `phasetype_decomposition_bayes.csv`; `S6_Fig`)

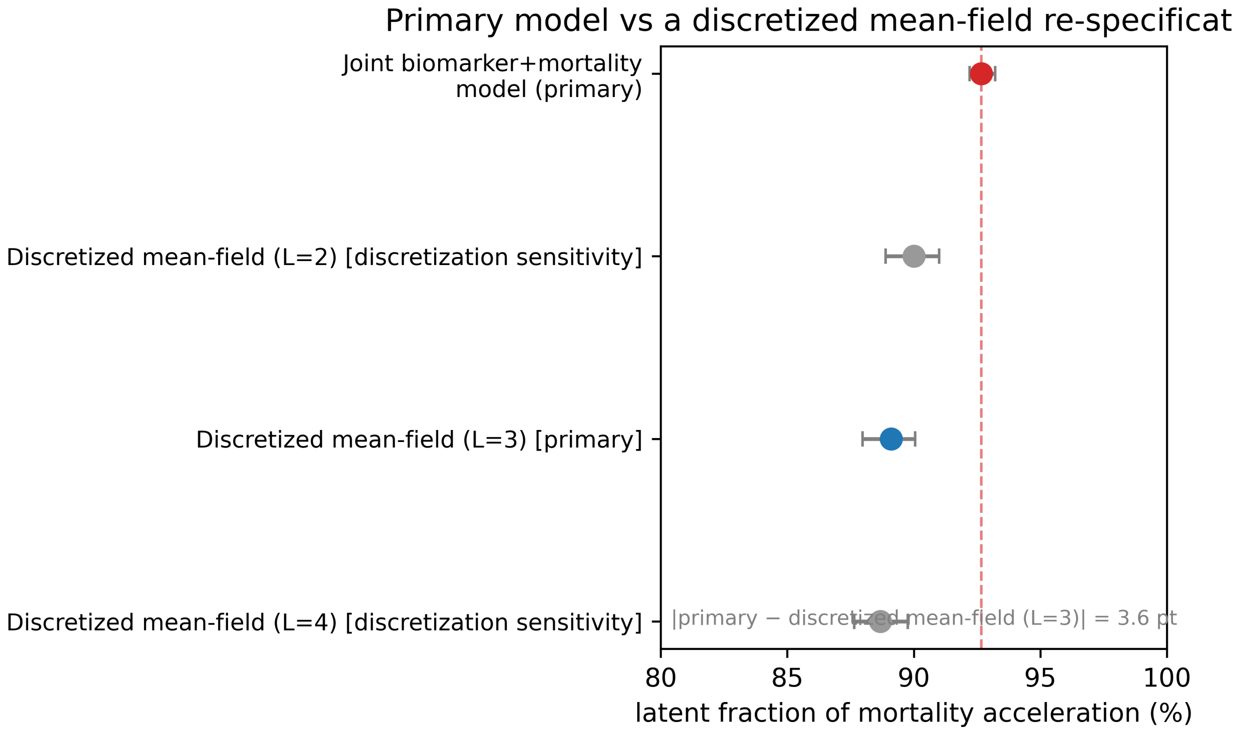

**Supplementary Table 1. Generator-matrix posterior (unified model).** Posterior summaries for the joint biomarker-and-mortality model. *Source:* NHANES 1999–2010 + Linked Mortality (n = 23,512). (`bayes_unified_summary.csv`, `bayes_unified_axis.csv`)

**Supplementary Table 2. HRS replication and blood-axis mediation.** Latent-fraction replication and Cox mediation by DNAm modules. *Source:* HRS (aggregate; n = 3,985, cell sizes ≥ 5). (`HRS_bayes_generator.csv`, `HRS_dnam_cox.csv`)

**Supplementary Table 3. DNAm module decomposition of the latent.** Reduction in latent fraction on adding each protein-trained DNAm module. *Source:* HRS (aggregate; n = 3,985, cell sizes ≥ 5). (`HRS_dnam_modules.csv`)

**Supplementary Table 4. Whole-blood RNA-seq module mortality hazard ratios.** *Source:* HRS (aggregate; n = 3,651, cell sizes ≥ 5). (`HRS_rnaseq_modules.csv`)

**Supplementary Table 5. Mendelian randomization, UKB-PPP.** Wald-ratio MR for latent components and known-causal positive controls. *Source:* UKB-PPP pQTL × parental-lifespan GWAS (GCST006697). (`MR_ukbppp_oriented.csv`)

**Supplementary Table 6. Mendelian randomization and colocalization, deCODE.** Cross-platform replication. *Source:* deCODE pQTL × parental-lifespan GWAS. (`MR_COLOC_decode_oriented.csv`)

**Supplementary Table 7. Mendelian randomization and colocalization, growth-signalling/IIS axis.** *Source:* UKB-PPP and deCODE pQTL × parental-lifespan GWAS. (`MR_COLOC_iis_oriented.csv`, `MR_COLOC_igf1_oriented.csv`)

**Supplementary Table 8. Colocalization, UKB-PPP.** Per-region coloc.abf posterior probabilities for all five hypotheses (H0 no association; H1 exposure only; H2 outcome only; H3 distinct causal variants; H4 shared causal variant), at the ±100 kb window with $p_1=p_2=10^{-4}$, $p_{12}=10^{-5}$. These columns are descriptive: neither positive control colocalized (LPA PP.H4 = 0.000, IL6R PP.H4 = 0.345), so a low PP.H4 is not used as evidence against causality; see Supplementary Table 17 for the prior/window sensitivity and Supplementary Methods §3. *Source:* UKB-PPP pQTL × parental-lifespan GWAS. (`COLOC_ukbppp.csv`)

**Supplementary Table 9. Reprogramming reversibility: donor-level and day-stratified clock outputs.** Per-donor Δ (reprogrammed − control) with 95% CI, n donors, one-sample t-test on donor means, and the day-stratified breakdown; the earlier pooled/unpaired values are retained alongside for comparison. *Source:* GEO GSE165179 (MPTR), GSE165178 (Sendai), GSE142439 (mRNA); public in-vitro reprogramming datasets. (`REV_GSE165179_donorlevel.csv`, `REV_GSE165178_donorlevel.csv`, `REV_damage.csv`, `REV_GSE165178.csv`, `REV_GSE142439.csv`)

**Supplementary Table 10. In-vivo mouse reprogramming (age-adjusted).** *Source:* GEO GSE190665 (public). (`TRACK2_mouse_clock_ageadj.csv`)

**Supplementary Table 11. Cross-species mortality-rate-doubling-time–lifespan anchor.** *Source:* AnAge database (public). (`S1_crossspecies_scaling.csv`)

**Supplementary Table 12. Minimum-detectable-effect (power) analysis of the MR nulls.** For each null protein, the minimum causal effect detectable at 80% power (two-sided α=0.05; MDE = 2.80 × SE), against the positive-control effects. `detects_LPA_scale` compares the MDE with the LPA effect (0.027 SD/SD) and `detects_IL6R_scale` with the larger of the two IL6R estimates (0.009 SD/SD [UKB-PPP], 0.015 SD/SD [deCODE]); no test reaches the smaller IL6R effect. *Source:* `mr_null_power.py` (from `MR_ukbppp_oriented.csv`, `MR_COLOC_iis_oriented.csv`, `MR_COLOC_decode_oriented.csv`).

**Supplementary Table 13. Posterior-predictive calibration by age band.** Observed versus posterior-predicted death counts (posterior-mean expected count and 95% predictive band) for the primary NHANES model. *Source:* NHANES 1999–2010 + Linked Mortality (n = 23,512). (`ppc_calibration.csv`)

**Supplementary Table 14. Subgroup robustness of the latent fraction.** Latent fraction with bootstrap 95% CI within sex, age group and NHANES cycle. *Source:* NHANES 1999–2010 + Linked Mortality (n = 23,512). (`stratified_latent.csv`)

**Supplementary Table 15. Prior sensitivity of the latent fraction (covariate-adjusted primary model).** Latent fraction, γ_base, r̂, bulk/tail ESS and divergences for three γ_base prior centres (N(0.04,0.03), N(0.08,0.03) primary, N(0.12,0.03)) on the covariate-adjusted model; all return 92.7% [92.2–93.2]. *Source:* NHANES 1999–2010 + Linked Mortality (n = 23,512). (`prior_sensitivity_covariate.py`; `prior_sensitivity_covariate.csv`)

**Supplementary Table 16. Identifiability and interval calibration of the latent-fraction estimator.** For a grid of known true latent fractions (60–95%): mean recovered value, bias, spread, and 95%-CI coverage over 40 simulated datasets each. Recovery is unbiased (|bias| ≤ 1.3 pt) and monotonic; mean coverage 0.93. *Source:* simulation (`identifiability_coverage.py`; `identifiability_coverage.csv`).

**Supplementary Table 17. Sensitivity of colocalization to the cis-window and the $p_{12}$ prior.** coloc.abf posteriors (H0–H4) for every combination of four cis-windows (±50, 100, 200, 500 kb) and five values of $p_{12}$ ($10^{-6}$, $5\times10^{-6}$, $10^{-5}$, $5\times10^{-5}$, $10^{-4}$), with $p_1=p_2=10^{-4}$ and $W=0.15^2$ held fixed; 100 cells. The published setting is (100 kb, $10^{-5}$). Whether the IL6R positive control colocalizes is set by the prior (PP.H4 0.047→0.843) and is insensitive to the window (≤0.005); LPA never colocalizes (PP.H3 ≥ 0.997); for CST3 and SERPINE1 the mass is on H1 (exposure only) throughout, and PP.H3 for SERPINE1 stays ≤ 0.082, so its nominal MR association is not attributable to linkage disequilibrium. *Source:* `coloc_sensitivity.py`. (`COLOC_sensitivity.csv`)

**Supplementary Table 18. Robustness of the causation-layer nulls (three sheets).** *Sheet `multiplicity`*: raw, Bonferroni-adjusted and Benjamini–Hochberg q values for the seven non-control markers tested on the primary platform. Neither of the two nominal associations (GHR p=0.018, SERPINE1 p=0.022) survives Bonferroni (α=0.05/7=0.007) or FDR control at 5% (smallest q=0.076); the family was fixed in advance and is not enlarged by the deCODE analyses. *Sheet `outcome_GWAS_sensitivity`*: the same lead-variant MR repeated against a larger parental-lifespan GWAS (Timmers et al. 2019, GCST009890, ≥500,193 offspring, log hazard protection ratios), with variants bridged GRCh38→rsID→GRCh37 through the harmonised GCST006697 file. Every non-control marker remains null and both nominal signals weaken (SERPINE1 p=0.15, GHR p=0.038); the outcome-side z-statistics show no gain in power (positive-control geometric mean ratio 0.91, and IL6R is detected less strongly), so Pilling 2017 is retained as the primary outcome. Timmers 2019 contains the Pilling UK Biobank sample, so this is a robustness check rather than independent replication. *Sheet `IGF1_fallback`*: direct IGF-1, whose cis lead variant is absent from the outcome GWAS, tested through the strongest covered cis variant under the pre-specified exposure-side rule. All three covered genome-wide-significant cis variants are rare (MAF 0.0037–0.0048), so no valid instrument exists for IGF-1 against this outcome GWAS and the resulting Wald ratio is reported only to show that the marker remains untested rather than null. The second aptamer (8406_17) has no genome-wide-significant cis variant at all. *Source:* `mr_multiplicity.py`, `outcome_gwas_comparison.py`, `mr_igf1_fallback.py`. (`mr_multiplicity.csv`, `OUTCOME_GWAS_comparison.csv`, `MR_igf1_fallback.csv`)

| Protein | Platform | Instrument | MR β (SD/SD) | $MDE_{80}$ (SD/SD) | Interpretation |
|---|---|---|---:|---:|---|
| GDF15 | UKB-PPP | F=1924 | -0.0033 | 0.023 | informative: LPA-scale effect would be detected |
| CST3 | UKB-PPP | F=1499 | -0.0017 | 0.023 | informative: LPA-scale effect would be detected |

| Protein | Platform | Instrument | MR β (SD/SD) | $MDE_{80}$ (SD/SD) | Interpretation |
|---|---|---|---|---|---|
| IGFBP3 | UKB-PPP | lead 837 | +0.0013 | 0.014 | informative: LPA-scale effect would be detected |
| GHR | UKB-PPP | lead 1398 | -0.0087 | 0.010 | informative: LPA-scale effect would be detected |
| GDF15 | deCODE | strong | +0.0039 | 0.018 | informative: LPA-scale effect would be detected |
| SERPINE1 | UKB-PPP | F=161 | -0.0546 | 0.067 | underpowered; not resolved by this design |
| IGFBP1 | UKB-PPP | lead 9.2 | +0.0827 | 0.138 | underpowered; not resolved by this design |
| IGF1R | UKB-PPP | lead 48.5 | -0.0207 | 0.060 | underpowered; not resolved by this design |
| CST3 | deCODE | $-\log_{10}P=138$ | +0.0017 | 0.030 | underpowered; not resolved by this design |

Key: 5/9 null tests could detect an LPA-scale effect (0.027 SD/SD) at 80% power and 2/9 the larger IL6R effect (0.015 SD/SD); none could exclude an effect as small as the smaller IL6R effect (0.009 SD/SD), which is not claimed.
